# The Construction and Analysis of ceRNA Network and Immune Infiltration in Kidney Renal Clear Cell Carcinoma

**DOI:** 10.1101/2021.02.07.21251289

**Authors:** Lugang Deng, Zhi Qu, Peixi Wang, Nan Liu

## Abstract

**Background:** Kidney renal clear cell carcinoma (KIRC) has the highest invasion, mortality and metastasis of the renal cell carcinomas and seriously affects patients’ quality of life. However, the composition of the immune microenvironment and regulatory mechanisms at transcriptomic level such as ceRNA of KIRC are still unclear.

**Methods:** We constructed a ceRNA network associated with KIRC by analyzing the long noncoding RNA (lncRNA), miRNA and mRNA expression data of 506 tumor tissue samples and 71 normal adjacent tissue samples downloaded from the Cancer Genome Atlas (TCGA) database. In addition, we estimated the proportion of 22 immune cell types in these samples through “The Cell Type Identification by Estimating Relative Subsets of RNA Transcripts”. Based on the ceRNA network and immune cells screened by univariate Cox analysis and Lasso regression, two nomograms were constructed to predict the prognosis of patients with KIRC. Receiver operating characteristic curves (ROC) and calibration curves were employed to assess the discrimination and accuracy of the nomograms. Consequently, co-expression analysis was carried out to explore the relationship between each prognostic gene in a Cox proportional hazards regression model of ceRNA and each survival-related immune cell in a Cox proportional hazards regression model of immune cell types to reveal the potential regulatory mechanism.

**Results:** We established a ceRNA network consisting of 12 lncRNAs, 25 miRNAs and 136 mRNAs. Two nomograms containing seven prognostic genes and two immune cells, respectively, were successfully constructed. Both ROC [Area Under Curves (AUCs) of 1, 3 and 5-year survival in the nomogram based on ceRNA network: 0.779, 0.747 and 0.772; AUCs of 1, 3 and 5-year survivals in nomogram based on immune cells: 0.603, 0.642 and 0.607] and calibration curves indicated good accuracy and clinical application value of both models. Through co-correlation analysis between ceRNA and immune cells, we found both LINC00894 and KIAA1324 were positively correlated with follicular helper T (Tfh) cells and negatively correlated with resting mast cells.

**Conclusions:** Based on the ceRNA network and tumor-infiltrating immune cells, we constructed two nomograms to predict the survival of KIRC patients and demonstrated their value in improving the personalized management of KIRC.

## Introduction

Renal cell carcinoma is a malignant kidney tumor that the American Cancer Society reports was diagnosed in 65,340 patients, and caused 14,970 deaths, in the United States during 2018 (1). It is a serious health problem worldwide, and the economic burden of the disease has steadily increased during the last century (2). Kidney renal clear cell carcinoma (KIRC) is the subtype of renal cell carcinoma with the highest invasiveness, mortality and metastasis rates (3); it is typically screened and diagnosed through computed tomography (CT), magnetic resonance imaging (MRI) and pathological section testing in the clinic (4). Several difficulties continue to make KIRC a challenge to treat, including its stealth in early stages, a lack of effective biomarkers, radiation risks and the high cost of diagnostic imaging required. Underlying biomarkers and molecular mechanisms for the prevention, diagnosis and prognosis of KIRC require further studies.

Long noncoding RNAs (lncRNAs) contain more than 200 nucleotides without the function of encoding proteins and have achieved attention recently because of their potential for exploring novel biomarkers in diseases and for elucidating mechanisms of biological processes (5, 6). Current research reveals that lncRNA plays an important role in the genesis and development of tumors through interactions with competing endogenous RNA (ceRNA) or by acting as microRNA sponges (7). Tumor-infiltrating immune cells are a major component of the tumor microenvironment and affect the clinical outcomes and pathological staging of multiple tumor types (8, 9). Some studies suggest lncRNA can affect tumor development and tumor immune cell microenvironment through the ceRNA network (10-12). Therefore, exploring interactions between the ceRNA network and various immune cells in the development of KIRC is essential.

In this study, we construct a ceRNA network related to the occurrence and development of KIRC using transcriptome and clinical data from the Cancer Genome Atlas (TCGA) to explore potential molecular mechanisms. In addition, we used “The Cell Type Identification by Estimating Relative Subsets of RNA Transcripts algorithm” (CIBERSORT) algorithm to assess differences in the composition of immune cells within tumor tissues and normal tissues. Furthermore, predictive nomograms were constructed based on the ceRNA network and on significant immune cells. Finally, we evaluated the relationship between tumor-infiltrating immune cells and the ceRNA network to identify the underlying regulatory mechanisms.

## Materials and methods

### Data source and selection

A flowchart illustrating the research process of our study is provided in Figure 1. Data on lncRNA, miRNA and mRNA expression of KIRC patients were downloaded from the Cancer Genome Atlas (TCGA) database (https://portal.gdc.cancer.gov/) through the GDC data transfer tool. Relevant clinical information including age, gender, survival data (vital status, days to death and days to last follow-up), grade and TNM stage of tumor tissues was also obtained for further analysis. We excluded samples with incomplete survival or pathological staging data, as well as duplicate samples. Only samples with both mRNA and miRNA expression profiles were included in this study. Correspondingly, the adjacent and paired normal tissues collected from the included patients was setted as the control group.

**Figure 1.**
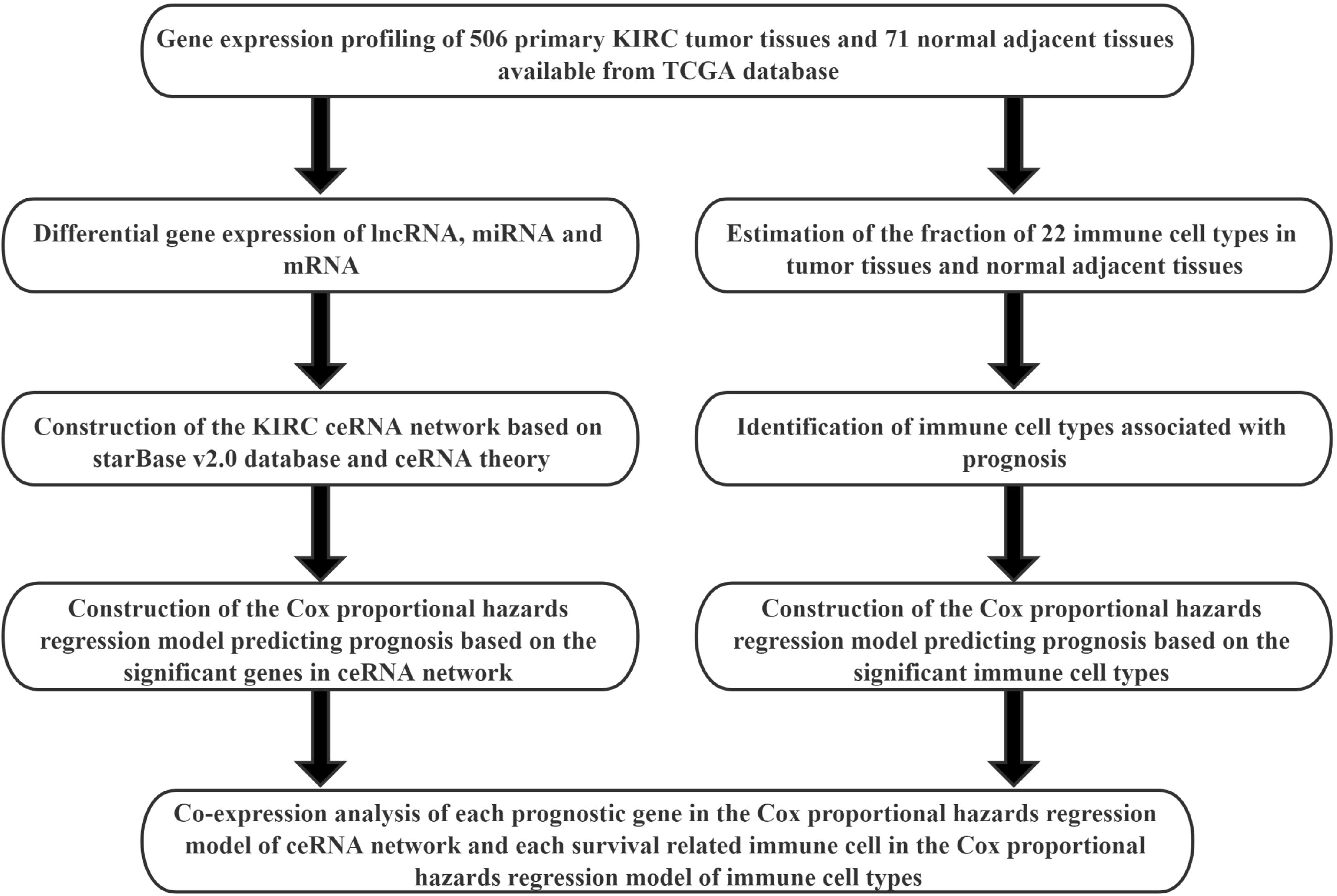
Flowchart of the analysis process.

### Differential expression analysis

Based on the HTSeq-Counts data of included samples, we used the “DESeq2” package (13) (http://bioconductor.org/packages/DESeq2/) of Bioconductor to screen for differentially expressed lncRNAs (DElncRNAs), miRNAs (DEmiRNAs) and mRNAs (DEmRNAs) between KIRC tissue and normal adjacent tissue. The log fold change (FC) criterion for differential expression was set as log_2_(FC) > 1 and the false discovery rate (FDR) at < 0.05. For the differential expression results of each type of RNA, volcano plots and heatmaps were generated through R packages “ggpolt2” and “pheatmap.”

### Construction of lncRNA-miRNA-mRNA ceRNA network and survival analysis

Using the differential expression results, we predicted the miRNAs-lncRNAs interactions and the miRNAs-mRNAs interactions based on the starBase v2.0 database (14, 15) (http://starbase.sysu.edu.cn/starbase2/index.php), which contained the miRNA-target interactions overlapped with CLIP-Seq data. We conducted hypergeometric tests and correlation analysis to filter the predicted interactions with FDR < 0.05 as the screening criteria. In addition, the lncRNA-miRNA-mRNA regulatory network of KIRC was visualized using Cytoscape v.3.8.1 (http://cytoscape.org/) (16). Besides, we conducted a Kaplan-Meier (K-M) survival curve analysis to assess the prognostic value of genes in the KIRC ceRNA network.

### ceRNA network: Cox proportional hazards regression model

Firstly, significant variables were screened for integration into the initial Cox model using univariate Cox analysis. Next, a least absolute shrinkage and selection operator (Lasso) regression was employed to further evaluate the fitness of multifaceted models and filter variables. Finally, we generated a nomogram of from multivariate Cox proportional hazards regression models to predict the prognosis of KIRC patients. To confirm the discrimination and precision of the nomogram, receiver operating characteristic curves (ROC) and calibration curves were employed. In addition, we calculated the risk score of each patient and then divided patients into high and low risk groups based on the median to conduct a K-M survival curve analysis.

### Tumor-infiltrating immune cells: CIBERSORT estimation and survival analysis

To analyze differences between the two groups at the level of immune cells, and explore the connection between key biomarkers in the ceRNA network and immune cells, we estimated the proportion of 22 kinds of immune cell types in 506 tumor tissues and 71 normal adjacent tissues by CIBERSORT (17) (http://cibersort.stanford.edu/), an algorithm for characterizing cell composition of complex tissues from their gene expression profiles. Only samples with CIBERSORT output of *P* < 0.05 were included in further analysis. After screening samples, we searched for immune cells with significant differences between cancer tissues and normal adjacent tissues, as indicated by a Wilcoxon rank-sum test. A K-M survival curve analysis was performed to evaluate the relationship between the content of different immune cells and the overall survival of KIRC patients.

### Tumor-infiltrating immune cells: Cox proportional hazards regression model

Immune cells with significant effects in univariate Cox analysis were integrated into the initial Cox model and the Lasso regression again used to evaluate the fitness of multifaceted models and filter variables further. As above, we constructed a nomogram based on the multivariate model of immune cells, and used ROC and calibration curves to confirm the discrimination and precision of the nomogram. Similarly, we conducted a K-M survival curve analysis by dividing patients into high and low risk groups according to the median of risk score. Finally, a Pearson correlation analysis was used to explore the relationship between genes and immune cells.

### Statistical Analysis

Only two-sided *P*-values < 0.05 was considered statistically significant. All statistical analyses were conducted in R version 4.0.2 software (Institute for Statistics and Mathematics, Vienna, Austria; www.r-project.org) (packages: corrplot, DESeq2, GDCRNATools, ggplot2, glmnet, pheatmap, rms, survival, survminer, timeROC).

## Results

### Differential expression in lncRNAs, miRNAs and mRNAs

After screening the samples of TCGA-KIRC, we constructed an experimental group and a control group containing of 506 tumor tissues and 71 normal adjacent tissues, respectively. Compared to the control group, we identified 357 DElncRNAs (287 up-regulated and 70 down-regulated), 132 DEmiRNAs (61 up-regulated and 71 down-regulated) and 3092 DEmRNAs (2032 up-regulated and 1060 down-regulated) using the cutoffs of the |log_2_(FC)| > 1 and FDR < 0.05 (Figure 2a-g).

**Figure 2.**
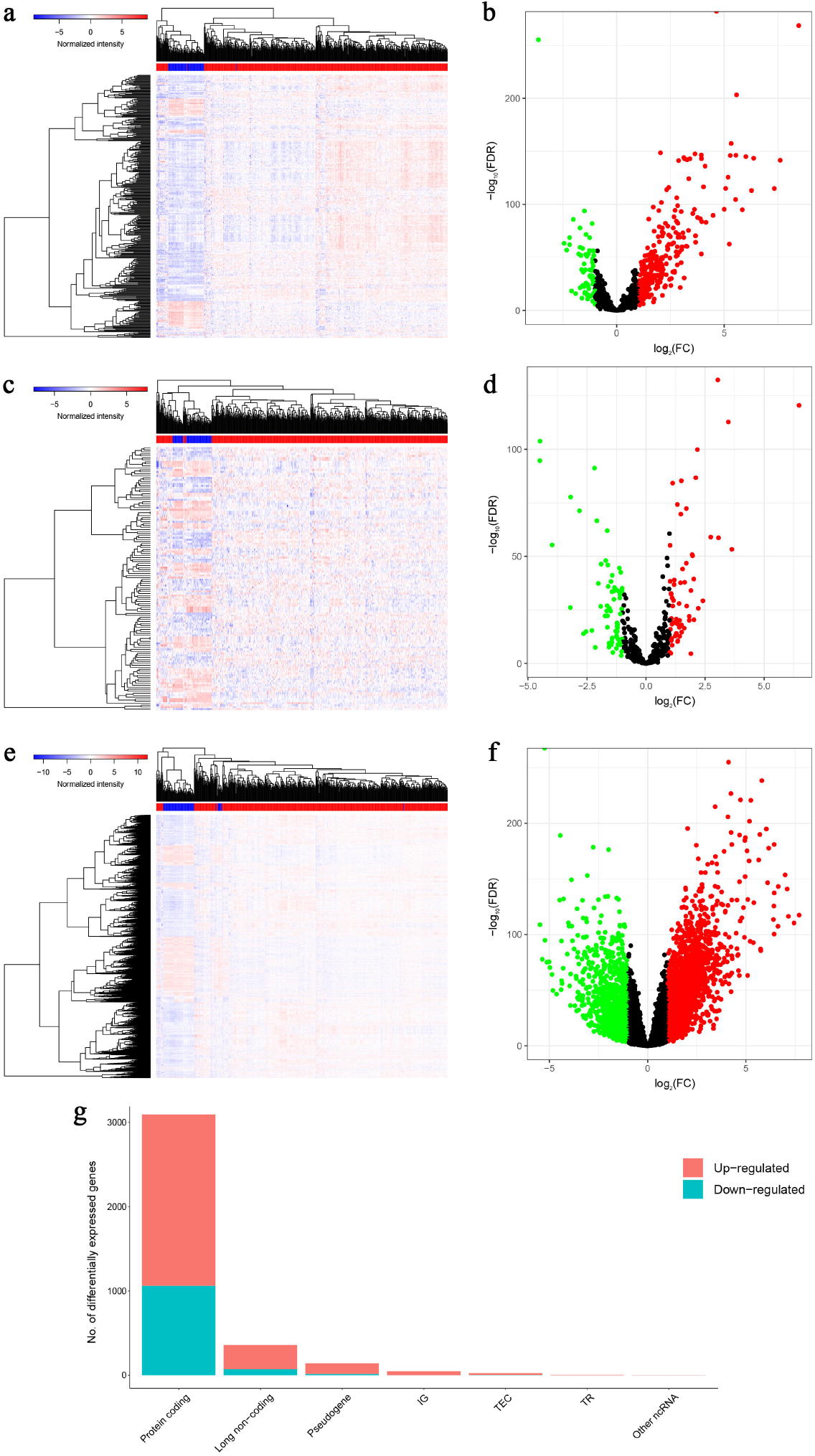
Genes differentially expressed in 506 KIRC tissues versus 71 normal adjacent tissues. Provided are the heatmap (a) and the volcano plot (b) of 357 differentially expressed lncRNAs between the two groups; the heatmap (c) and the volcano plot (d) of 132 differentially expressed miRNAs between the two groups; and the heatmap (e) and the volcano plot (f) of 3092 differentially expressed protein-coding genes between the two groups; and the composition of differentially expressed genes (g). All differentially expressed genes had a log (fold-change) > 1.0 or < −1.0 and the overall of FDR < 0.05.

### Construction of lncRNA-miRNA-mRNA ceRNA network and survival analysis

Using the starBase v2.0 database and ceRNA theory, a lncRNA-miRNA-mRNA ceRNA network consisting of 173 genes (12 lncRNAs, 25 miRNAs and 136 mRNAs) was established (Figure 3a and Table 1). A K-M survival curve analysis to assess the prognostic value of nodes in the ceRNA network revealed 83 genes (10 lncRNAs, 7 miRNAs and 66 mRNAs) related to the overall survival of KIRC patients. Survival curves of the top three (according to *P*-value) lncRNAs (PVT1, AC005154.1, AC015813.1), miRNAs (hsa-miR-21-5p, hsa-miR-130b-3p, hsa-miR-204-5p), and mRNAs (MXD3, VPS13D, KCNN4) are shown in Figures 3b-d, e-g and h-j, respectively.

**Table 1.** Hypergeometric testing and correlation analysis results of ceRNAs pairs.

| lncRNAs | mRNAs | miRNAs | Hypergeometric test *P* | Correlation *P* |
| --- | --- | --- | --- | --- |
| AC005154.1 | KAT2A | hsa-miR-122-5p | 0.042851557 | 1.32E-95 |
| AC015813.1 | SLC16A1 | hsa-miR-590-3p | 0.0433213 | 0.019445968 |
| AC015813.1 | FHOD1 | hsa-miR-590-3p | 0.007220217 | 4.09E-19 |
| AC015813.1 | E2F8 | hsa-miR-590-3p | 0.025270758 | 1.22E-09 |
| AC015813.1 | SERPINB9 | hsa-miR-590-3p | 0.025270758 | 0.000636067 |
| AC015813.1 | COL5A2 | hsa-miR-590-3p | 0.014440433 | 0.015398545 |
| AC015813.1 | BRIP1 | hsa-miR-590-3p | 0.003610108 | 5.18E-09 |
| AC015813.1 | ESCO2 | hsa-miR-590-3p | 0.02166065 | 0.009163956 |
| AC015813.1 | TXLNB | hsa-miR-590-3p | 0.003610108 | 0.020621823 |
| AC015813.1 | PSTPIP2 | hsa-miR-590-3p | 0.003610108 | 0.000108913 |
| AC015813.1 | SNRNP70 | hsa-miR-590-3p | 0.003610108 | 4.80E-73 |
| AC016876.2 | TNFRSF10B | hsa-miR-133a-3p | 0.003810813 | 9.33E-39 |
| AC016876.2 | MYO9B | hsa-miR-133a-3p | 0.000392403 | 1.06E-53 |
| AC016876.2 | PREX1 | hsa-miR-133a-3p | 0.013191013 | 2.17E-22 |
| AC016876.2 | CAPN15 | hsa-miR-133a-3p | 0.027474351 | 1.51E-34 |
| AC016876.2 | CORO1C | hsa-miR-133a-3p | 0.001461641 | 2.06E-36 |
| AC016876.2 | FJX1 | hsa-miR-133a-3p | 0.036258784 | 6.66E-19 |
| AC016876.2 | NDRG1 | hsa-miR-133a-3p | 0.010361047 | 1.00E-11 |
| AC016876.2 | VKORC1 | hsa-miR-133a-3p | 0.001165794 | 1.01E-27 |
| AC016876.2 | TMEM200B | hsa-miR-133a-3p | 0.013191013 | 9.12E-19 |
| AC016876.2 | RSRP1 | hsa-miR-133a-3p | 0.000881161 | 6.23E-07 |
| C1RL-AS1 | SCARB1 | hsa-miR-16-5p | 0.001438811 | 1.64E-42 |
| C1RL-AS1 | VEGFA | hsa-miR-16-5p | 0.030764401 | 2.81E-61 |
| C1RL-AS1 | AHNAK2 | hsa-miR-16-5p | 0.000392403 | 7.17E-21 |
| C1RL-AS1 | PTHLH | hsa-miR-16-5p | 0.009888558 | 1.10E-07 |
| C1RL-AS1 | RELT | hsa-miR-16-5p | 0.000732486 | 3.61E-14 |
| C1RL-AS1 | PFKFB4 | hsa-miR-16-5p | 0.004002511 | 6.80E-27 |
| C1RL-AS1 | CCND1 | hsa-miR-16-5p | 0.046303563 | 7.81E-25 |
| C1RL-AS1 | APLN | hsa-miR-16-5p | 0.000549364 | 2.74E-25 |
| C1RL-AS1 | SH2D2A | hsa-miR-16-5p | 0.000732486 | 7.13E-19 |
| C1RL-AS1 | SEL1L3 | hsa-miR-16-5p | 0.012975462 | 1.23E-23 |
| C1RL-AS1 | UNC5B | hsa-miR-16-5p | 0.000392403 | 1.58E-15 |
| C1RL-AS1 | PHLDA3 | hsa-miR-16-5p | 0.001177209 | 0.000714118 |
| C1RL-AS1 | SLC41A2 | hsa-miR-16-5p | 0.000941767 | 1.16E-10 |
| C1RL-AS1 | GRB10 | hsa-miR-16-5p | 0.022523937 | 4.08E-21 |
| C1RL-AS1 | SLC2A3 | hsa-miR-16-5p | 0.013812588 | 1.39E-09 |
| C1RL-AS1 | MYBL1 | hsa-miR-16-5p | 0.049468948 | 1.21E-08 |
| C1RL-AS1 | ANLN | hsa-miR-16-5p | 0.004002511 | 7.40E-05 |
| C1RL-AS1 | OTOGL | hsa-miR-16-5p | 0.000392403 | 1.38E-08 |
| C1RL-AS1 | KCNJ2 | hsa-miR-16-5p | 0.019384712 | 9.72E-07 |
| C1RL-AS1 | CNTNAP1 | hsa-miR-16-5p | 0.003139225 | 6.23E-12 |
| C1RL-AS1 | NRARP | hsa-miR-16-5p | 0.023622665 | 4.17E-13 |
| C1RL-AS1 | CREB5 | hsa-miR-16-5p | 0.013812588 | 5.44E-48 |
| C1RL-AS1 | ATAD5 | hsa-miR-16-5p | 0.042928897 | 2.56E-37 |
| C1RL-AS1 | INSR | hsa-miR-16-5p | 0.009182232 | 1.03E-20 |
| C1RL-AS1 | SIPA1L2 | hsa-miR-16-5p | 0.006043007 | 9.08E-08 |
| C1RL-AS1 | AATK | hsa-miR-16-5p | 0.000549364 | 1.52E-28 |
| C1RL-AS1 | RFLNB | hsa-miR-16-5p | 0.000941767 | 5.34E-05 |
| C1RL-AS1 | SPRY4 | hsa-miR-16-5p | 0.002746822 | 8.52E-14 |
| C1RL-AS1 | ARL10 | hsa-miR-16-5p | 0.001438811 | 1.17E-11 |
| C1RL-AS1 | ANKRD13B | hsa-miR-16-5p | 0.003139225 | 7.32E-16 |
| C1RL-AS1 | SNAP25 | hsa-miR-16-5p | 0.002380579 | 1.30E-19 |
| C1RL-AS1 | LY6E | hsa-miR-16-5p | 0.000941767 | 1.68E-17 |
| C1RL-AS1 | CDK5R1 | hsa-miR-16-5p | 0.013812588 | 1.69E-16 |
| C1RL-AS1 | CLCN5 | hsa-miR-16-5p | 0.006618532 | 0.020236846 |
| C1RL-AS1 | BDNF | hsa-miR-16-5p | 0.010621043 | 8.30E-09 |
| C1RL-AS1 | IRX3 | hsa-miR-16-5p | 0.000732486 | 1.79E-37 |
| C1RL-AS1 | PLAGL1 | hsa-miR-16-5p | 0.004473395 | 1.30E-23 |
| C1RL-AS1 | IRF4 | hsa-miR-16-5p | 0.004970439 | 2.67E-12 |
| C1RL-AS1 | TSPYL2 | hsa-miR-16-5p | 0.002380579 | 1.01E-20 |
| C1RL-AS1 | TLL1 | hsa-miR-16-5p | 0.007848062 | 5.19E-19 |
| C1RL-AS1 | KCNN4 | hsa-miR-16-5p | 0.000732486 | 4.96E-09 |
| C1RL-AS1 | TRIM36 | hsa-miR-16-5p | 0.015565322 | 0.000352309 |
| C1RL-AS1 | CBFA2T3 | hsa-miR-16-5p | 0.004970439 | 6.86E-13 |
| C1RL-AS1 | TSC22D3 | hsa-miR-16-5p | 0.012975462 | 3.07E-05 |
| C1RL-AS1 | SNCG | hsa-miR-16-5p | 0.001726574 | 0.02336615 |
| EPB41L4A-AS1 | TNFRSF10B | hsa-miR-93-5p | 0.005296055 | 0.000309792 |
| EPB41L4A-AS1 | IKBIP | hsa-miR-93-5p | 0.049992152 | 1.07E-06 |
| H19 | EGLN3 | hsa-miR-130b-3p | 0.025364366 | 0.030923622 |
| H19 | UXS1 | hsa-miR-130b-3p | 0.017341521 | 0.000663666 |
| H19 | USP46 | hsa-miR-130b-3p | 0.024521918 | 0.043104031 |
| H19 | ACSL4 | hsa-miR-130b-3p | 0.049356887 | 0.046472711 |
| H19 | LDLRAD3 | hsa-miR-130b-3p | 0.017341521 | 2.91E-06 |
| H19 | BMP6 | hsa-miR-130b-3p | 0.021136246 | 1.44E-05 |
| H19 | RASD1 | hsa-miR-130b-3p | 0.001092495 | 0.000237995 |
| H19 | NHSL1 | hsa-miR-130b-3p | 0.002162624 | 0.000635296 |
| H19 | C8orf4 | hsa-miR-130b-3p | 0.040677495 | 1.83E-10 |
| H19 | MAP3K12 | hsa-miR-130b-3p | 0.000287237 | 0.006322847 |
| H19 | VPS13D | hsa-miR-130b-3p | 0.00310273 | 0.022110203 |
| H19 | PXDN | hsa-miR-130b-3p | 0.001415171 | 2.64E-12 |
| H19 | PMEPA1 | hsa-miR-130b-3p | 0.001092495 | 2.21E-10 |
| H19 | WNK3 | hsa-miR-130b-3p | 0.000207327 | 0.000409846 |
| H19 | PFKFB3 | hsa-miR-130b-3p | 0.004199543 | 0.001757017 |
| H19 | TCF4 | hsa-miR-130b-3p | 0.000309855 | 2.83E-07 |
| H19 | HECW2 | hsa-miR-130b-3p | 0.001092495 | 0.000515485 |
| H19 | LRRC41 | hsa-miR-130b-3p | 0.008478302 | 0.004641957 |
| H19 | KLHL3 | hsa-miR-130b-3p | 0.035134428 | 0.004796761 |
| H19 | LDLR | hsa-miR-130b-3p | 0.002491825 | 6.36E-13 |
| H19 | SCARA3 | hsa-miR-130b-3p | 0.005054152 | 6.50E-06 |
| LINC00893 | SCARB1 | hsa-miR-125b-5p, hsa-miR-125a-5p | 0.001438811 | 0.000105621 |
| LINC00893 | VEGFA | hsa-miR-125b-5p, hsa-miR-125a-5p | 0.030764401 | 8.10E-26 |
| LINC00893 | MYEOV | hsa-miR-125b-5p | 0.014414273 | 6.39E-05 |
| LINC00893 | SOX11 | hsa-miR-125b-5p, hsa-miR-125a-5p | 0.015565322 | 9.42E-07 |
| LINC00893 | PRDM1 | hsa-miR-125b-5p, hsa-miR-125a-5p | 0.032046251 | 0.001791829 |
| LINC00893 | GJC1 | hsa-miR-125b-5p, hsa-miR-125a-5p | 0.003139225 | 1.58E-05 |
| LINC00893 | OLFML2A | hsa-miR-125b-5p, hsa-miR-125a-5p | 0.000156961 | 0.009879398 |
| LINC00893 | LIMD2 | hsa-miR-125b-5p, hsa-miR-125a-5p | 0.006043007 | 8.66E-13 |
| LINC00893 | GRB10 | hsa-miR-125b-5p, hsa-miR-125a-5p | 0.022523937 | 0.004617551 |
| LINC00893 | TNFAIP3 | hsa-miR-125b-5p, hsa-miR-125a-5p | 0.02040496 | 0.000292635 |
| LINC00893 | KMT5C | hsa-miR-125b-5p, hsa-miR-125a-5p | 0.006618532 | 1.47E-81 |
| LINC00893 | PDE7A | hsa-miR-125b-5p, hsa-miR-125a-5p | 0.019384712 | 2.94E-13 |
| LINC00893 | PLAGL1 | hsa-miR-125b-5p, hsa-miR-125a-5p | 0.004473395 | 1.50E-09 |
| LINC00893 | IRF4 | hsa-miR-125b-5p, hsa-miR-125a-5p | 0.004970439 | 8.51E-05 |
| LINC00893 | FAM118A | hsa-miR-125b-5p, hsa-miR-125a-5p | 0.005493643 | 1.23E-43 |
| LINC00894 | GRIN2D | hsa-miR-342-3p | 0.02166065 | 0.020258812 |
| MALAT1 | ZNF395 | hsa-miR-429, hsa-miR-200c-3p | 0.033565927 | 0.009006143 |
| MALAT1 | SCD | hsa-miR-200b-3p, hsa-miR-200a-3p, hsa-miR-429,<br>hsa-miR-200c-3p, hsa-miR-141-3p | 0.039289444 | 0.011807116 |
| MALAT1 | EZH2 | hsa-miR-217 | 0.039289444 | 6.86E-14 |
| MALAT1 | MXD3 | hsa-miR-200b-3p, hsa-miR-429, hsa-miR-200c-3p | 0.001707738 | 3.90E-34 |
| MALAT1 | BAZ1A | hsa-miR-590-3p | 0.018390826 | 7.26E-05 |
| MALAT1 | SIX1 | hsa-miR-200b-3p, hsa-miR-200a-3p, hsa-miR-429,<br>hsa-miR-200c-3p, hsa-miR-141-3p, hsa-miR-590-3p | 0.00025153 | 4.87E-06 |
| MALAT1 | KCNJ2 | hsa-miR-1-3p, hsa-miR-1271-5p, hsa-miR-590-3p | 0.000680978 | 0.01227854 |
| MALAT1 | RUNX2 | hsa-miR-203a-3p, hsa-miR-217, hsa-miR-590-3p | 0.027070651 | 0.007921587 |
| MALAT1 | PDE7A | hsa-miR-1-3p | 0.031465278 | 1.64E-07 |
| MALAT1 | CCND2 | hsa-miR-200a-3p, hsa-miR-141-3p, hsa-miR-1-3p, hsa-miR-1271-5p | 0.002367103 | 0.005749454 |
| MALAT1 | FOXF1 | hsa-miR-200b-3p, hsa-miR-429, hsa-miR-200c-3p | 0.033565927 | 0.001701238 |
| MALAT1 | KIAA1324 | hsa-miR-1271-5p | 0.040612149 | 0.018453328 |
| NEAT1 | STC2 | hsa-miR-181b-5p, hsa-miR-204-5p | 0.005508592 | 0.022782701 |
| NEAT1 | GJC1 | hsa-let-7e-5p, hsa-let-7g-5p | 0.010508709 | 0.038550157 |
| NEAT1 | ADM | hsa-miR-181b-5p | 0.028501136 | 9.33E-08 |
| NEAT1 | EZH2 | hsa-let-7e-5p, hsa-let-7g-5p | 0.008654278 | 1.27E-12 |
| NEAT1 | AURKB | hsa-let-7e-5p, hsa-let-7g-5p | 0.00048449 | 0.032101352 |
| NEAT1 | KIF21B | hsa-let-7e-5p, hsa-let-7g-5p | 0.003786827 | 3.94E-06 |
| NEAT1 | INTS6L | hsa-let-7e-5p, hsa-let-7g-5p | 0.001348287 | 1.00E-46 |
| NEAT1 | DTX2 | hsa-let-7e-5p, hsa-let-7g-5p | 0.022864398 | 0.003873252 |
| NEAT1 | LIMD2 | hsa-let-7e-5p, hsa-let-7g-5p | 0.001852057 | 0.023584192 |
| NEAT1 | NGF | hsa-let-7e-5p, hsa-let-7g-5p | 0.00048449 | 0.036481826 |
| NEAT1 | POLQ | hsa-miR-181b-5p, hsa-let-7e-5p | 0.026346413 | 4.13E-05 |
| NEAT1 | POU2F2 | hsa-let-7e-5p, hsa-let-7g-5p | 0.00772659 | 0.046579228 |
| NEAT1 | APBB3 | hsa-let-7e-5p, hsa-let-7g-5p | 0.003786827 | 1.03E-93 |
| NEAT1 | MAPK11 | hsa-let-7e-5p, hsa-let-7g-5p | 0.00048449 | 0.020903367 |
| NEAT1 | FBXO41 | hsa-miR-181b-5p | 0.000291346 | 0.00718285 |
| NEAT1 | PITPNM3 | hsa-let-7e-5p, hsa-let-7g-5p | 0.022864398 | 0.000826877 |
| NEAT1 | NPHP3 | hsa-let-7e-5p, hsa-let-7g-5p | 0.022864398 | 7.31E-68 |
| NEAT1 | DNA2 | hsa-let-7e-5p, hsa-let-7g-5p | 0.003786827 | 1.69E-34 |
| NEAT1 | AMT | hsa-let-7e-5p, hsa-let-7g-5p | 0.000906364 | 0.001503922 |
| NEAT1 | SLC25A37 | hsa-miR-181b-5p | 0.002491825 | 1.48E-28 |
| NEAT1 | FAM118A | hsa-let-7e-5p, hsa-let-7g-5p | 0.034971322 | 7.22E-34 |
| NEAT1 | AASS | hsa-let-7e-5p, hsa-let-7g-5p | 0.00048449 | 1.39E-05 |
| NEAT1 | CELSR3 | hsa-miR-181b-5p, hsa-miR-204-5p | 0.00026878 | 5.77E-21 |
| NEAT1 | KIFC2 | hsa-let-7e-5p, hsa-let-7g-5p | 0.002491825 | 4.59E-65 |
| NEAT1 | RNF165 | hsa-let-7e-5p, hsa-let-7g-5p | 0.00587435 | 2.76E-05 |
| NEAT1 | SLC25A27 | hsa-let-7e-5p, hsa-let-7g-5p | 0.001554187 | 2.36E-89 |
| PVT1 | TNFRSF10B | hsa-miR-93-5p | 0.005296055 | 1.15E-70 |
| PVT1 | IKBIP | hsa-miR-93-5p | 0.049992152 | 4.69E-68 |
| PVT1 | TET3 | hsa-miR-93-5p, hsa-miR-20b-5p | 0.000351901 | 6.58E-37 |
| PVT1 | CCND2 | hsa-miR-93-5p, hsa-miR-20b-5p | 0.000180512 | 2.67E-22 |
| SNHG1 | NETO2 | hsa-miR-21-5p | 0.041029046 | 3.64E-18 |
| SNHG1 | CDKN2C | hsa-miR-154-5p | 0.02166065 | 9.52E-06 |
| SNHG1 | GBP1 | hsa-miR-21-5p | 0.02166065 | 0.014315805 |
| SNHG1 | ADGRG2 | hsa-miR-21-5p | 0.00064624 | 1.35E-05 |

**Figure 3.**
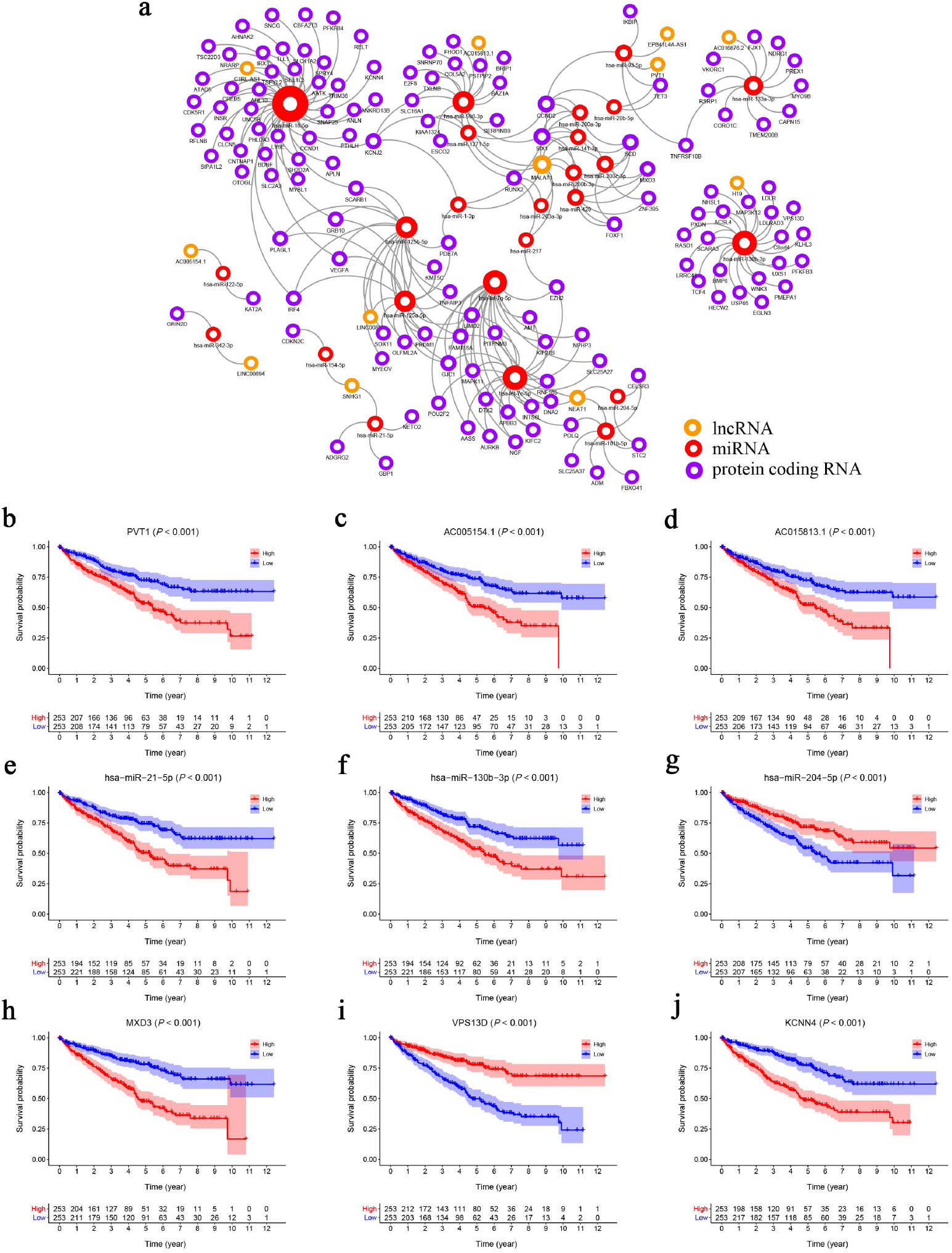
(a) The KIRC related ceRNA network consisting of 12 lncRNAs, 25 miRNAs and 136 mRNAs. (b-j) The K-M survival curves of the top three lncRNAs, miRNAs and mRNAs: PVT1 (b), AC005154.1 (c), AC015813.1 (d), hsa-miR-21-5p (e), hsa-miR-130b-3p (f), hsa-miR-204-5p (g), MXD3 (h), VPS13D (i) and KCNN4 (j).

### ceRNA network: Cox proportional hazards regression model

Univariate Cox analysis applied to filter genes in the ceRNA network retained 97 genes for incorporation into the initial model (Table S1). Lasso regression analysis indicated that 15 genes were statistically significant in this model. Subsequently, a Cox proportional hazards regression model with seven genes was established through further screening of key genes based on the Akaike Information Criterion (AIC). Finally, we constructed a nomogram to predict the 1, 3 and 5-year overall survival probability of KIRC patients. ROC [Area Under Curves (AUCs)] for the model gave 1, 3 and 5-year survivals of 0.779, 0.747 and 0.772, respectively, which reflected the discrimination and precision of the nomogram with calibration curves (Figure 4). Overall survival (based on K-M curves) of the high-risk group is lower than that of the low-risk group (Figure S1). Survival curves of the genes in the model are also provided in Figure S1.

**Figure 4.**
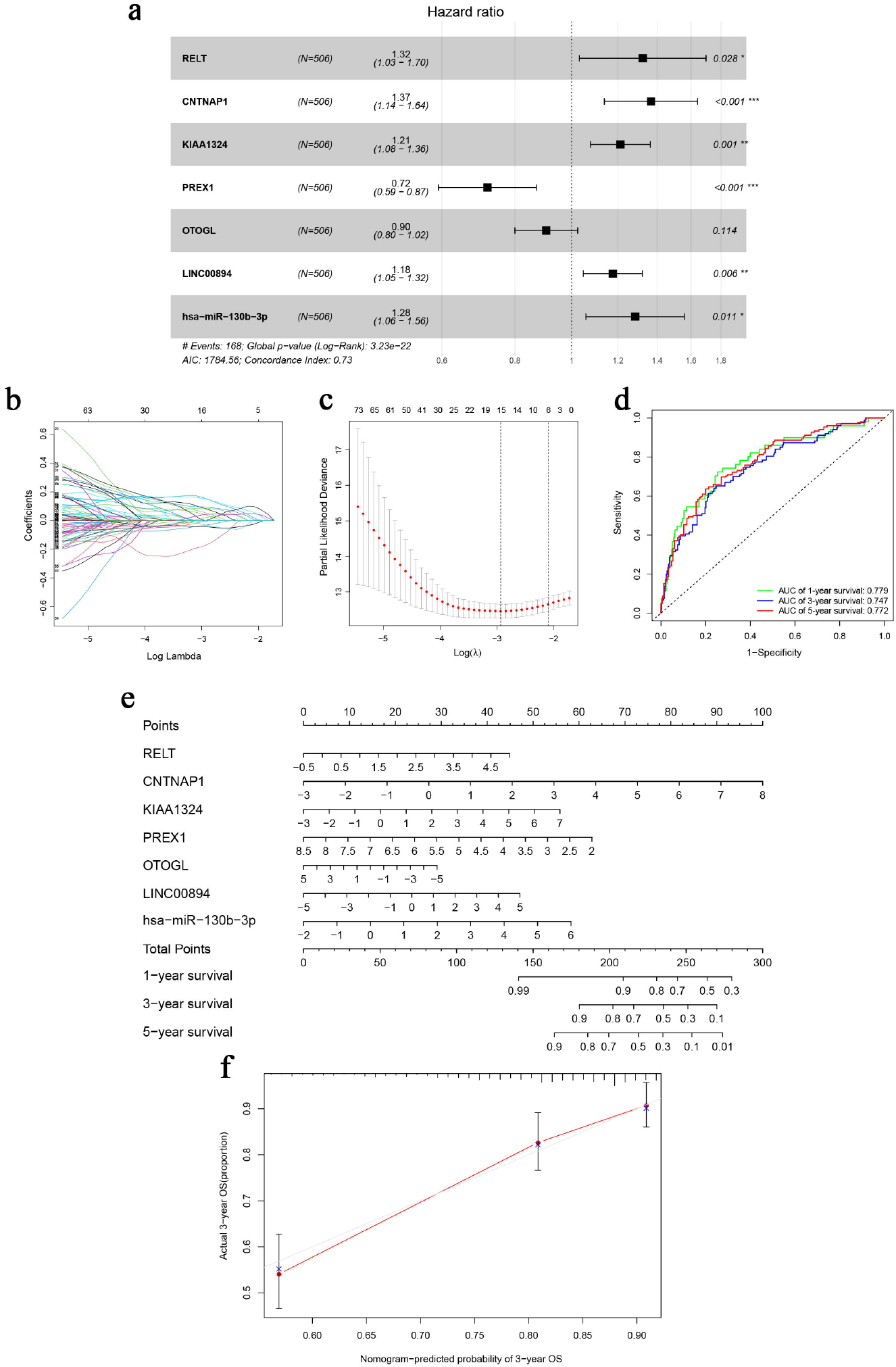
The Cox proportional hazards regression model (a) based on RNAs screened by Lasso regression analysis (b and c) in the ceRNA network. Seven potential prognosis related RNAs were integrated into the Cox proportional hazards regression model. The nomogram (e) was constructed based on the model. The ROC (d) and calibration curves (f) indicate the acceptable accuracy (AUCs of 1, 3 and 5-year survivals: 0.779, 0.747 and 0.772) and discrimination of the nomogram. \**P* < 0.05; \*\**P* < 0.010; \*\*\**P* < 0.001.

### Tumor-infiltrating immune cells: CIBERSORT estimation and survival analysis

The analysis by CIBERSORT identified five normal tissue samples and 206 tumor tissue samples for inclusion in subsequent analysis (*P <* 0.05) and the proportions of 22 immune cells in each sample are displayed in the histogram and heat map of Figure 5. A Wilcoxon rank-sum test showed that naive B cells (*P* < 0.001), plasma cells (*P* < 0.001), CD8 T cells (*P* = 0.014), CD4 naive T cells (*P* < 0.001), CD4 memory resting T cells (*P* = 0.013), regulatory (Tregs) T cells (*P* = 0.015), gamma delta T cells (*P* = 0.021), resting dendritic cells (*P* = 0.016) and resting mast cells (*P* = 0.048) all varied between the two tissue sample types (Figure 5c). Based on the results of K-M survival curve analysis, five immune cells, including memory B cells, plasma cells, follicular helper T (Tfh) cells, T regulatory (Tregs) cells and resting mast cells were related to overall survival of KIRC patients (Figure S2).The clinical correlation of 22 immune cells is displayed in Figure S3.

### Tumor-infiltrating immune cells: Cox proportional hazards regression model

After filtering based on univariate Cox analyses (Table S2), two of 22 immune cells (Tfh and resting mast cells) were incorporated into the initial regression model. Lasso regression analysis and AIC were then employed for further optimization of the model, and both cells were kept in the final Cox proportional hazards regression model. A nomogram was developed to predict the 1, 3 and 5-year overall survival probability of KIRC patients based on the model. Both the ROC (AUCs of 1, 3 and 5-year survivals: 0.603, 0.642 and 0.607) and the calibration curves suggested that the nomogram had good accuracy (Figure 6). The K-M survival curves suggested that significant differences existed in the overall survival of the high versus low-risk group (Figure S4).

**Figure 5.**
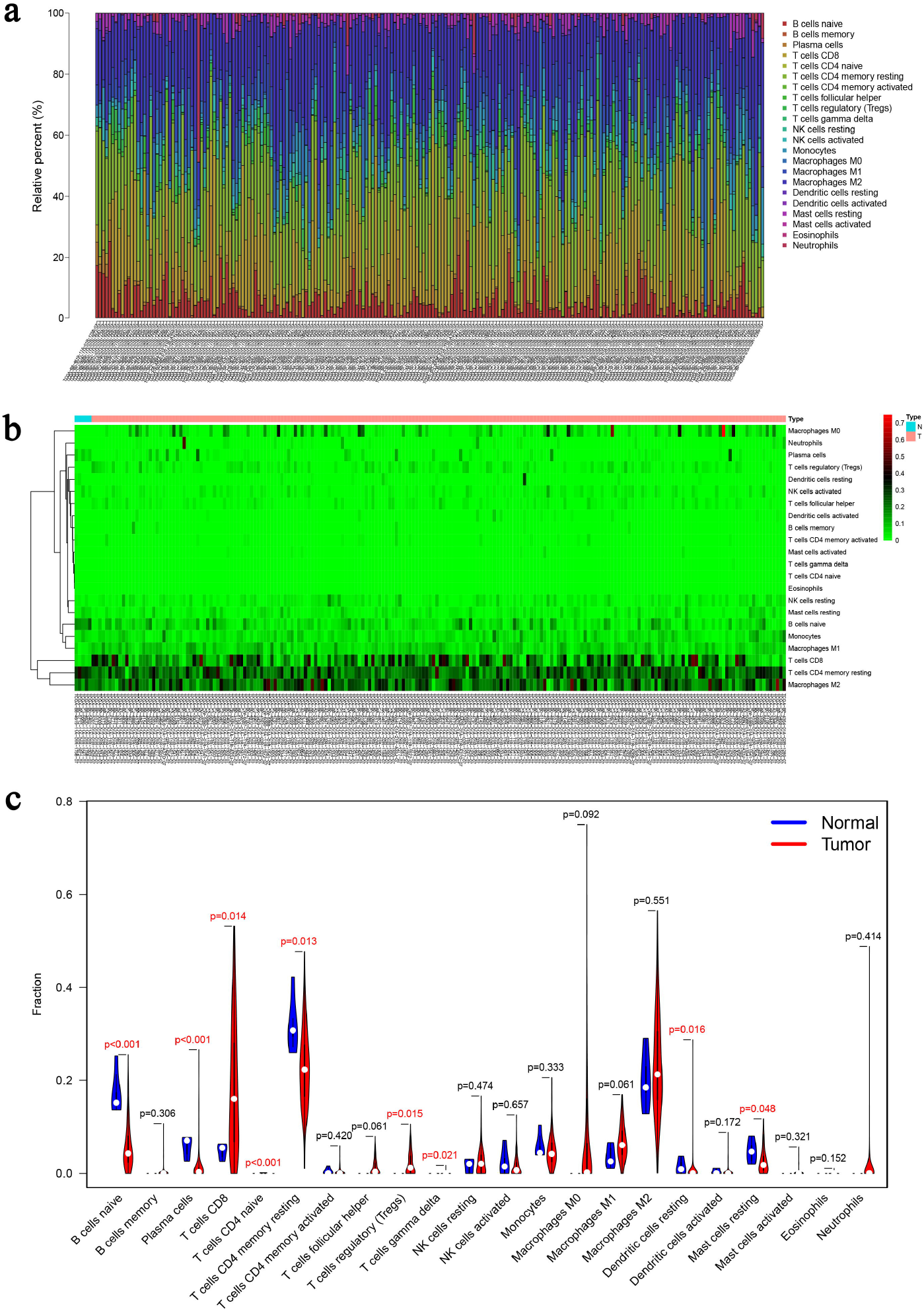
The composition (a) and heatmap (b) of immune cells estimated by the CIBERSORT algorithm in five normal adjacent tissues and 206 KIRC tissues. The violin plot of immune cells (c) compares cells’ proportion between the two groups.

**Figure 6.**
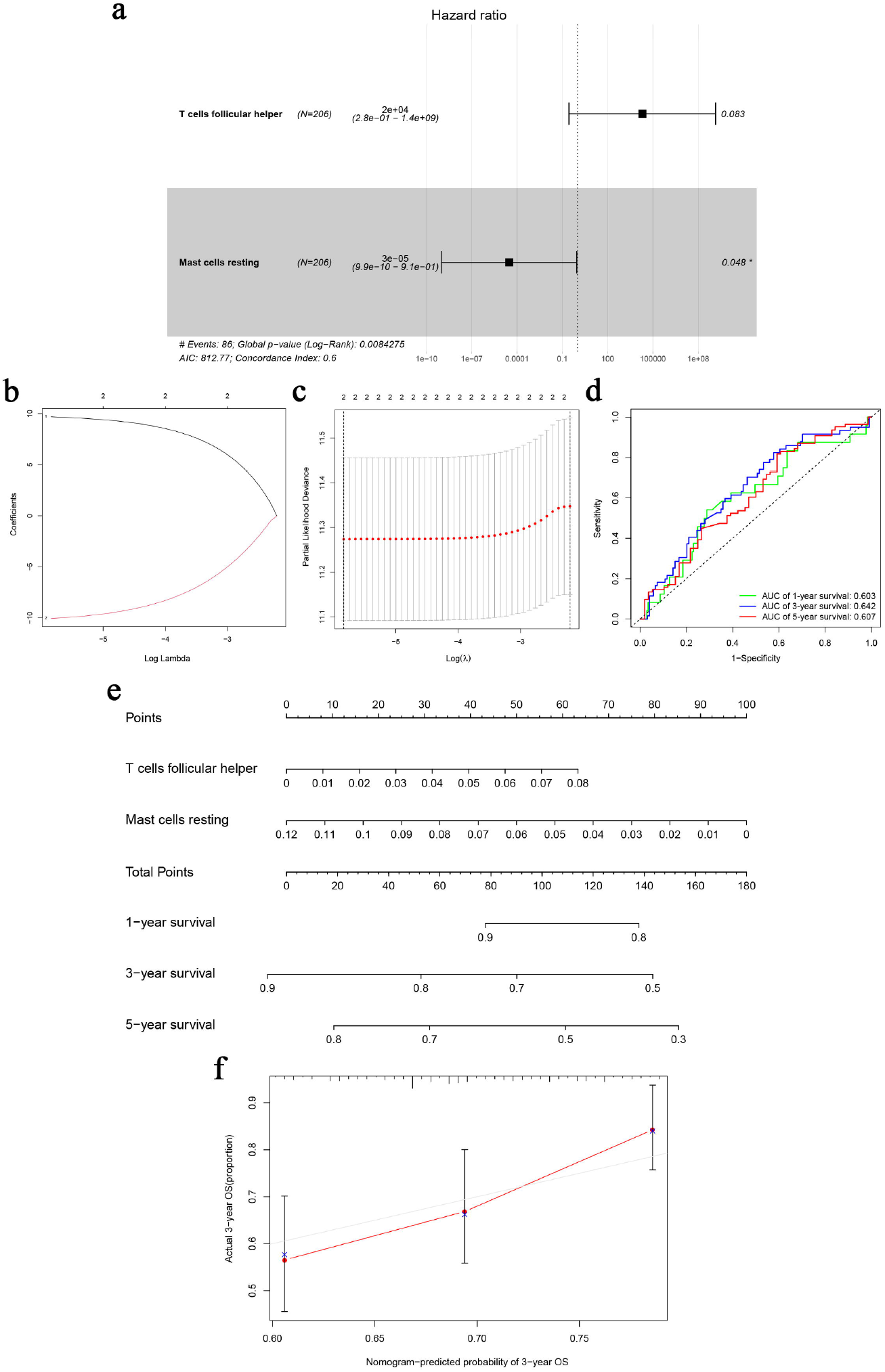
The Cox proportional hazards regression model (a) based on immune cells screened by Lasso regression analysis (b and c). Tfh cells and resting mast cells were integrated into the Cox proportional hazards regression model. The nomogram (e) was constructed based on the model. The ROC (d) and the calibration curves (f) indicate the acceptable accuracy (AUCs of 1, 3 and 5-year survivals: 0.603, 0.642 and 0.607) and discrimination of the nomogram. \**P* < 0.05; \*\**P* < 0.010; \*\*\**P* < 0.001.

### Co-expression analysis of key genes and immune cells

Pearson correlation analysis was used to explore the co-expression pattern between 22 types of immune cells (Figure 7a). Similarly, the co-expression relationship between the key biomarkers and immune cells of the two models is illustrated in Figure 7b. Tfh cells are positively correlated with LINC00894 (Figure 7c, R = 0.36, *P* < 0.001) and KIAA1324 (Figure 7d, R = 0.39, *P* < 0.001), while resting mast cells are negatively correlated with LINC00894 (Figure 7e, R = −0.32, *P* < 0.001) and KIAA1324 (Figure 7f, R = −0.30, *P* < 0.001).

**Figure 7.**
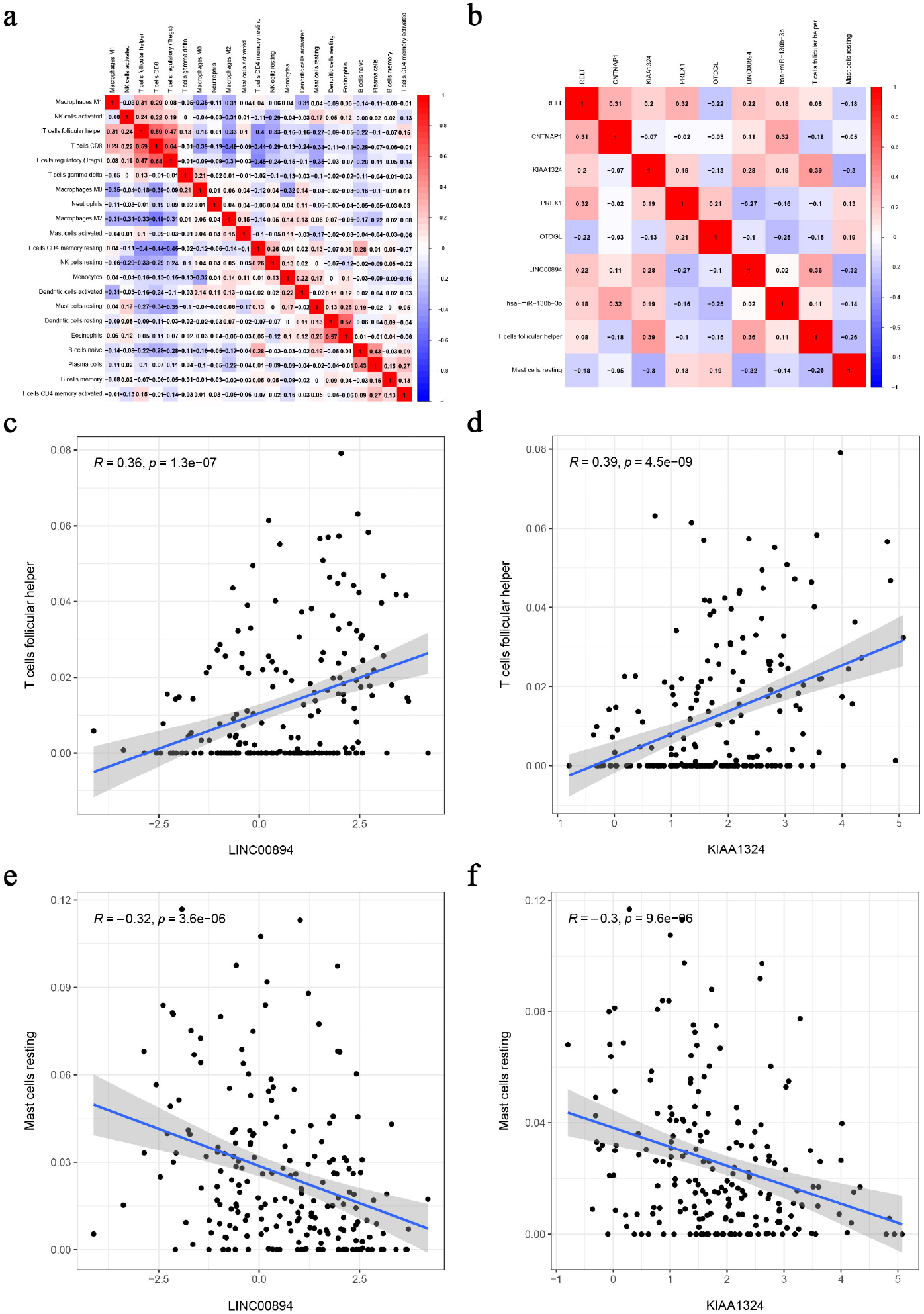
Co-expression heatmap of all immune cells in KIRC (a); the co-expression heatmap of the RNAs and immune cells in the two Cox proportional hazards regression models (b); the significant results of Pearson correlation coefficients between the RNAs and immune cells: Tfh cells and LINC00894 (c); Tfh cells and KIAA1324 (d); resting mast cells and LINC00894 (e); resting mast cells and KIAA1324 (f).

### Validation of the correlation between key genes and immune cells

Multiple databases were searched to verify the correlation between the key genes (LINC00894 and KIAA1324) and immune cells (Tfh cells and resting mast cells) identified. B cell lymphoma 6 (BCL6), CXCL13, CD10 (MME), ICOS and PD-1 (PDCD1) were the most common surface markers of Tfh cells in the CellMarker database (Figure S5). In the Oncomine(tm) platform, we explored the expression of KIAA1324 and Tfh markers among various cancers. KIAA1324, BCL6, CXCL13, ICOS and PDCD1 were highly expressed and MME was downregulated in KIRC compared to normal kidney tissue (Figure S6); median rank and *P*-values are listed in Table S3.

Using the Gene Expression Profiling Interactive Analysis (GEPIA), KIAA1324 is positively correlated with CXCL13, ICOS and PDCD1 in KIRC (Figure S7), and LINC00894 is positively correlated with BCL6, ICOS and PDCD1 in KIRC (Figure S8). Similarly, the results of the LinkedOmics database showed that KIAA1324 was positively correlated with BCL6, CXCL13, ICOS and PDCD1 in KIRC. In addition, the expression of KIAA1324 was significantly different among tumor stages and related to tumor purity in KIRC (Figure S9).

## Discussion

KIRC is one of the most common and malignant subtypes of kidney tumors (3). However, because of limitations in early detection and a lack of sensitive biomarkers, patients with KIRC typically have a poor prognosis. Recently, ceRNA was identified as playing a potentially important role in the molecular regulatory function of tumorigenesis and growth, and in affecting differences in immune-infiltrating cells within multiple tumors. In this study, we investigated the role and connection of the ceRNA network and immune infiltration in KIRC to explore key prognostic biomarkers and potential regulatory mechanisms.

We established a ceRNA network consisting of 12 lncRNAs, 25 miRNAs and 136 mRNAs and compared the composition differences of immune cells related to KIRC. Based on the results, two prediction nomograms consisting of seven genes and two immune cells, respectively, were constructed to evaluate overall survival. A correlation analysis of the key factors in both nomograms indicated that both LINC00894 (lncRNA) and KIAA1324 (protein-coding RNA) were positively correlated with Tfh cells and negatively correlated with resting mast cells. Consequently, we infer that ceRNA regulatory function, in which both LINC00894 and KIAA1324 participate, and Tfh cells and resting mast cells may play a crucial role in the development and treatment of KIRC.

LINC00894 is lncRNA derived from a locus on the X chromosome that is differentially expressed in various cancers (18). Similar to our work, Liang (19) reported that LINC00894 was a tumor-special lncRNA in KIRC that was upregulated in tumor tissues and correlated with overall survival time. Epithelial-to-mesenchymal transition (EMT) regulating miRNAs regulate drug resistance and cancer progression and metastasis (20, 21). LINC00894 can inhibit the TGF-β2/ZEB1 signaling pathway by acting as the sponge of EMT-regulating miR-200, or can be upregulated by ERα activation and positively regulate the expression of miR-200a-3p and miR-200b-3p, which results in induction of the TGF-β2/ZEB1 signaling pathway to reduce the occurrence of drug resistance in breast cancers (18, 22, 23). In our study, LINC00894 might suppress the expression of hsa-miR-342-3p, which has an important role in progression, staging and metastasis of various cancers (24-27). In view of the upregulation of hsa-miR-342-3p contributing to gefitinib resistance via targeting CPA4 in non-small cell lung cancer (28), we speculate that LINC00894 is involved in the progression of cancer and may be a promising treatment target to eliminate drug resistance in patients with KIRC.

KIAA1324, also known as EIG121, encodes a 1013 amino acid transmembrane protein that shows high sequence conservation among different species. In type I endometrial cancer (estrogen-related), the expression of KIAA1324 is upregulated, but it is downregulated in type II endometrial cancer (not estrogen-related), which is more malignant than type I (29). KIAA1324 may be a novel suppressor, being epigenetically downregulated in gastric cancer (30); in addition, its high expression in endometrial and pancreatic carcinoma predicts improved survival (31, 32). However, increased expression of KIAA1324 in ovarian cancer is associated with a poor prognosis (33). These studies indicate that KIAA1324 may exhibit different biological functions in various cancers. Furthermore, recent mechanistic studies reveal that KIAA1324 is localized to endosome-lysosome compartments and associated with autophagy, which helps protect cells from cell death by upregulating the autophagy pathway under unfavorable conditions, such as starvation or chemotherapy (34). Therefore, we infer that overexpressed KIAA1324 helps cancer cells proliferate and resist chemotherapy in KIRC.

In this study, we explored five among 22 types of immune cells related to patient prognosis, and screened out the Tfh cells and resting mast cells to construct a Cox proportional hazards regression model, which evaluation of its AUC suggests has clear clinical value. Gou et al. (35) also explored the relationship between immune cells and renal cell carcinoma, and similar to our research, found Tfh cells were associated with poor prognosis and resting mast cells were positively associated with long term survival.

Under the action of myeloid professional antigen presenting cells (APC), naive CD4+ T cells could differentiate into Tfh cells and non-Tfh effector cells (such as Th1, Th2, or Th17 cells) (36). Therefore, Tfh cells are a specialized subset of CD4+ T cells that help B cells respond to antigens (37), which is, in turn, depends on the expression of transcriptional factor B cell lymphoma 6 (BCL6) (38). In the past decade, several studies have revealed a potential connection between Tfh cells and infiltrated tumors. Amé-Thomas et al. (39) demonstrated that Tfh cells protect follicular lymphoma malignant B cells from spontaneous and rituximab-induced apoptosis by upregulating CD40L and IL-4, which might lead to a worse prognosis. In non-small cell lung cancer (NSCLC) tumor tissues, the proportion of Tfh cells is increased, suggesting that Tfh cells might play an important role in estimating the poor survival of NSCLC patients (40). In addition, Tfh cell abundance is known to improve prognosis of patients with breast cancer or colorectal cancer (41, 42), which indicates the value of Tfh cells against tumors through immune responses.

Mast cells, both resting and activated, are large granulated innate immune cells found predominantly in sites between the host and its external environment, such as skin, respiratory mucosa, and the gastrointestinal tract (43). The activation of mast cells is related to various stimuli received by numerous receptors on the cell surface, such as pathogens, neuropeptides, cytokines, growth factors, toxins, basic compounds, complement, immune complexes, etc. (44). Recent studies have shown that the association between mast cells and cancer is two-sided, including both involvement in, and protection against, tumor progression. Xu et al. (45) constructed a prognosis-associated microRNA–mast cell network in lung adenocarcinoma, and found resting mast cell numbers were closely related to better overall survival and disease-free survival (DFS), while activated mast cells were related to poor survival. miR-30a and miR-550a are also involved in the regulation of immune response by acting as the promoter and suppressor of resting mast cells, respectively. In contrast, mast cells in hepatocellular carcinoma are mostly resting, inactivation of mast cells can promote immune escape, which is beneficial to tumor growth (46).

Our research inevitably has some limitations that should be recognized. First, our research data was obtained from public databases. The limited data means that the clinical pathological parameters are incomplete, which can lead to potential errors and biases. Secondly, we have not taken the heterogeneity of the immune microenvironment associated with the location of immune infiltration into consideration. In other words, the accuracy and applicability of the prediction models will be affected by the heterogeneity of tissue subtypes. In addition, the data series used in this study are all from Western countries, so applying the research conclusions to Asian countries has certain bias and may be inappropriate. As well, the lack of cell markers for resting mast cells leads to incomplete validation in our study. Lastly, this study is not a biological mechanism research, as it lacks experiments on the interaction mechanisms between ceRNA and immune cells. Despite limitations of the study, we did combine tumor infiltrating immune cells with a ceRNA network for analysis in KIRC, which not only constructed two normograms with clinical predictive value, but also identified a correlation between two key prognostic molecules and two prognosis-related immune cells, providing novel directions for future research.

## Conclusions

Based on tumor-infiltrating immune cells and ceRNA networks, we constructed two nomograms to predict survival of KIRC patients. The high AUC values we obtained reflect the utility of this approach, which provides clinicians with more comprehensive clinical information through the models to improve individual management of KIRC patients. In addition, our study suggests that LINC00894, KIAA1324, Tfh cells and resting mast cells are significantly related to the prognosis of KIRC patients and we infer that LINC00894 and KIAA1324 might interact with Tfh cells and resting mast cells to affect the progression of KIRC. This study also provides new ideas for the pathogenesis and clinical treatment of KIRC.

## Data Availability

The dataset (TCGA-KIRC) analyzed in this study is publicly available in TCGA (https://tcga-data.nci.nih.gov/tcga/).The other data used to support the findings of this study are available from the corresponding author upon request.

https://tcga-data.nci.nih.gov/tcga/

## Author contribution

ZQ and NL conceived the idea and designed the study. LD conducted the data analysis and visualization. PW performed the data interpretation and literature collection. LD and NL wrote the manuscript. ZQ and NL contributed to the revision of the manuscript draft. All authors read and approved the final manuscript.

## Data availability statement

The dataset (TCGA-KIRC) analyzed in this study is publicly available in TCGA (https://tcga-data.nci.nih.gov/tcga/).

## Acknowledgement

This work was supported by National Natural Science Foundation of China (No. 81872584) and (No. 81472941), National 863 Young Scientist Program (No. 2015AA020940), Key Scientific Research Project Plan of Henan Province (No. 21A330001), Natural Science Foundation of Guangdong Province (No. 2016A030313138), Key Projects of Guangzhou Science and Technology Program (No. 201704020056), Interdisciplinary Research for First-class Discipline Construction Project of Henan University (No. 2019YLXKJC04) and Scientific Research Project for University of Education Bureau of Guangzhou (No. 201831841).

