## Supplementary File for "The Construction and Analysis of ceRNA Network and Immune Infiltration in Kidney Renal Clear Cell Carcinoma"

Lugang Deng ^1^, Peixi Wang ^2^, Zhi Qu ^2^, Nan Liu ^1, 2, 3, *^
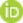

^1^ School of Public Health, Guangzhou Medical University, Guangzhou, 511436, P. R. China

^2^ Institute of Chronic Disease Risks Assessment, School of Nursing and Health, Henan University, Kaifeng, 475004, P. R. China

^3^ Pinghu Hospital, Health Science Center, Shenzhen University, Shenzhen, 518116, P. R. China.

*Corresponding authors:

**Nan Liu**

orcid.org/0000-0002-8895-3169

**Supporting Figures**

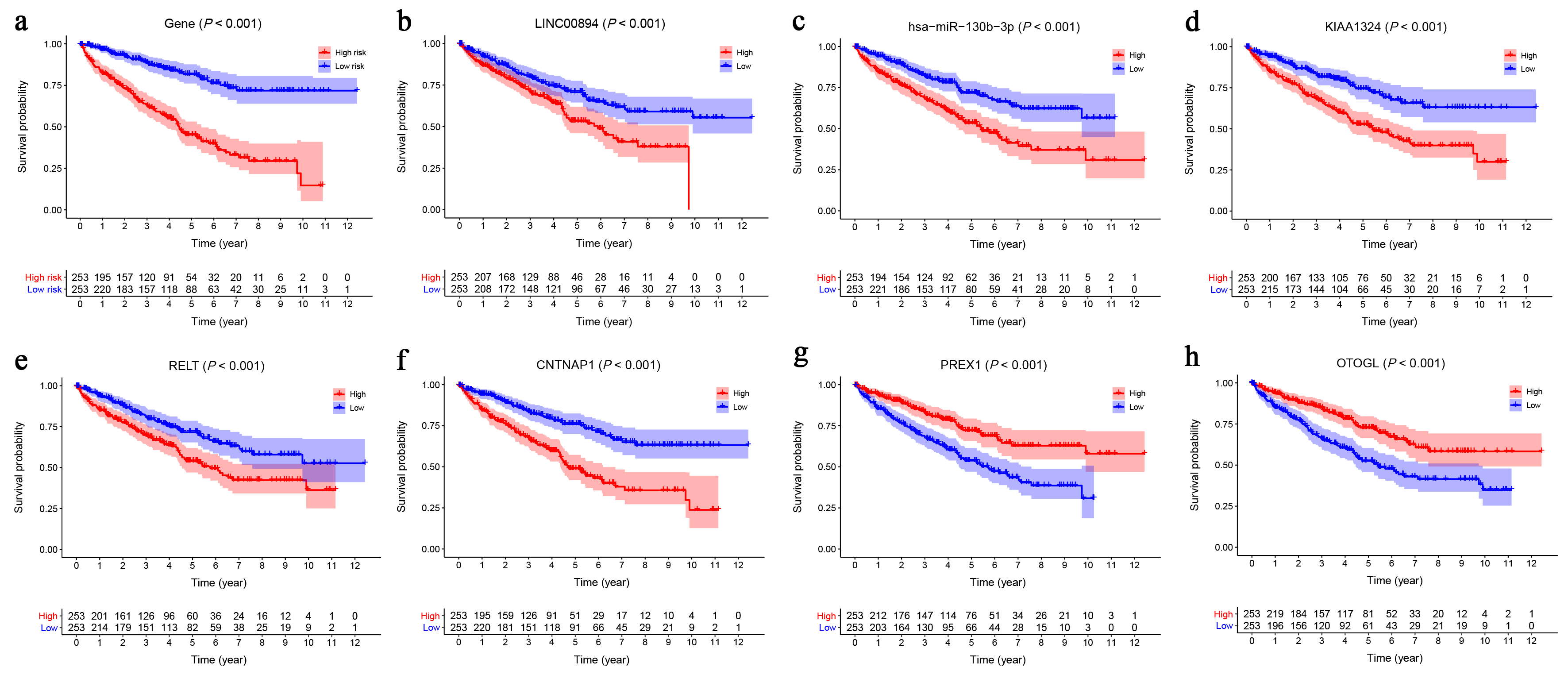

**Figure S1** The Kaplan-Meier survival analysis for examination of the association between the model based on the ceRNA network and prognosis.

(a) All the genes in the model based on the ceRNA network, (b) LINC00894, (c) hsa−miR−130b−3p, (d) KIAA1324, (e) RELT, (f) CNTNAP1, (g) PREX1 and (h) OTOGL.

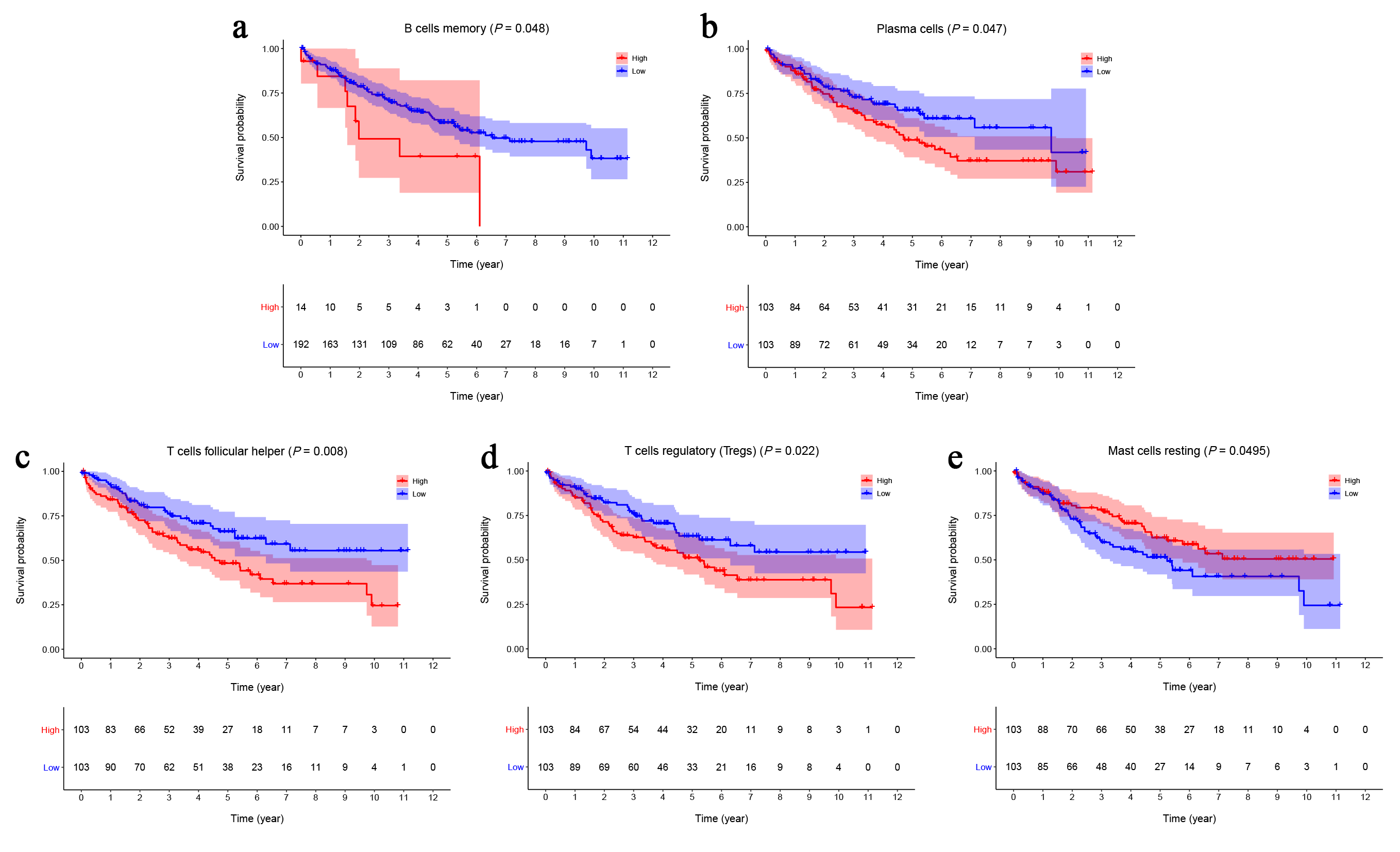

**Figure S2** The Kaplan-Meier survival curves of the immune cells related to the overall survival of KIRC patients.

(a) B cells memory, (b) Plasma cells, (c) Tfh cells, (d) T cells regulatory (Tregs) and (e) Mast cells resting.

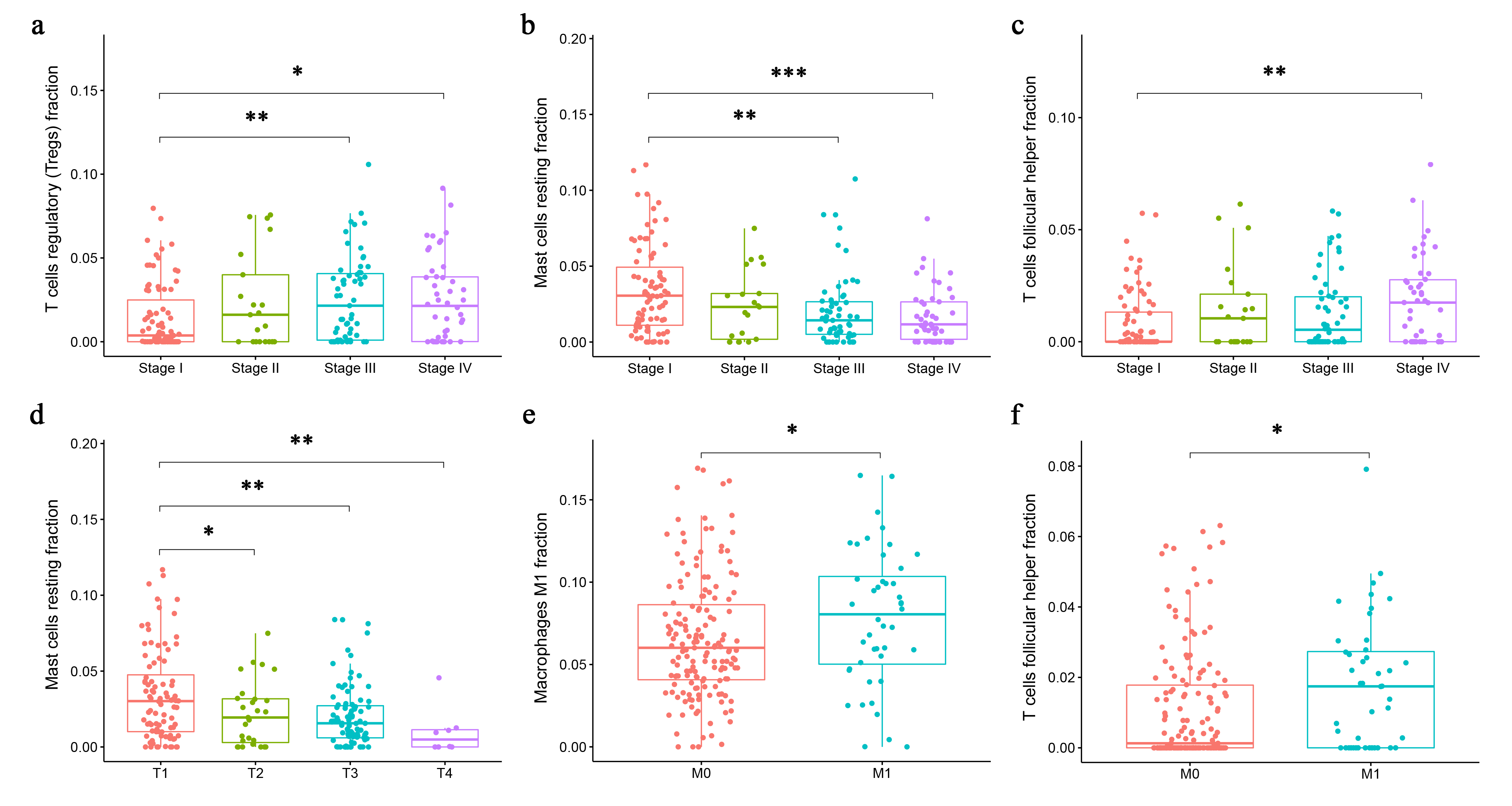

**Figure S3** The clinical correlation analysis of immune cells.

(a) The fraction of T regulatory (Tregs) cells in stage Ⅰ significantly differed from stage Ⅲ (*P* = 0.003) and stage Ⅳ (*P* = 0.015) with an increasing trend; (b) The fraction of resting mast cells in stage Ⅰ significantly differed from stage Ⅲ (*P* = 0.001) and stage Ⅳ (*P* < 0.001) with an declining trend; (c) The fraction of Tfh cells in stage Ⅳ was higher than stage Ⅰ (*P* = 0.002); (d) The fraction of resting mast cells in T1 stage significantly differed from T2 (*P* = 0.049), T3 (*P* = 0.001) and T4 stage (*P* = 0.009) by an declining trend; (e) The fraction of macrophages M1 in M1 stage was higher than M0 stage (*P* = 0.045); (f) The fraction of Tfh cells in M0 stage was lower than M1 stage (*P* = 0.012).

**P* < 0.05; ***P* < 0.01; ****P* < 0.001.

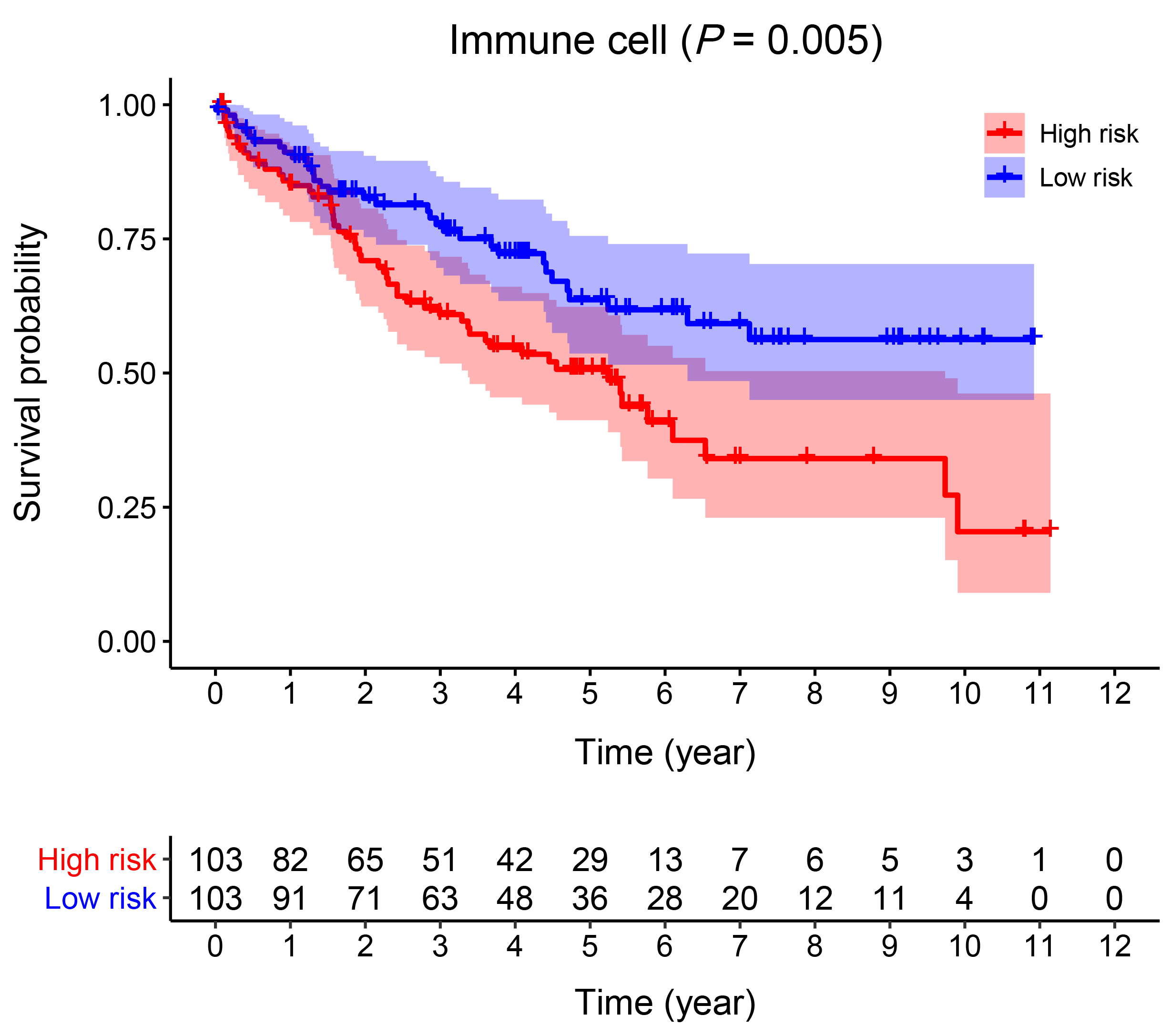

**Figure S4** Examination of the association between the model based on the immune cells and prognosis by Kaplan-Meier survival analysis.

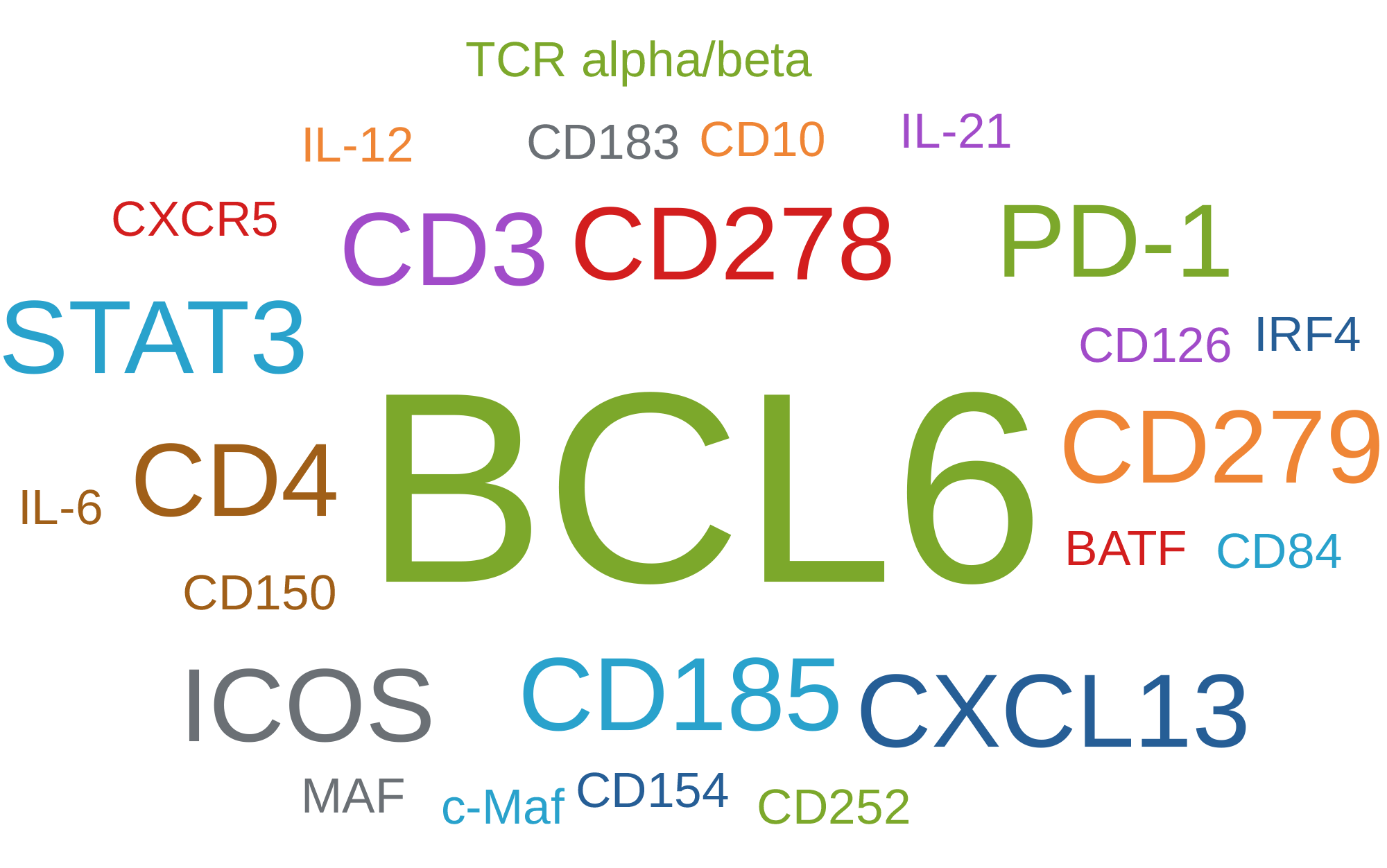

**Figure S5** The surface markers of the Tfh cells reported by CellMarker database.

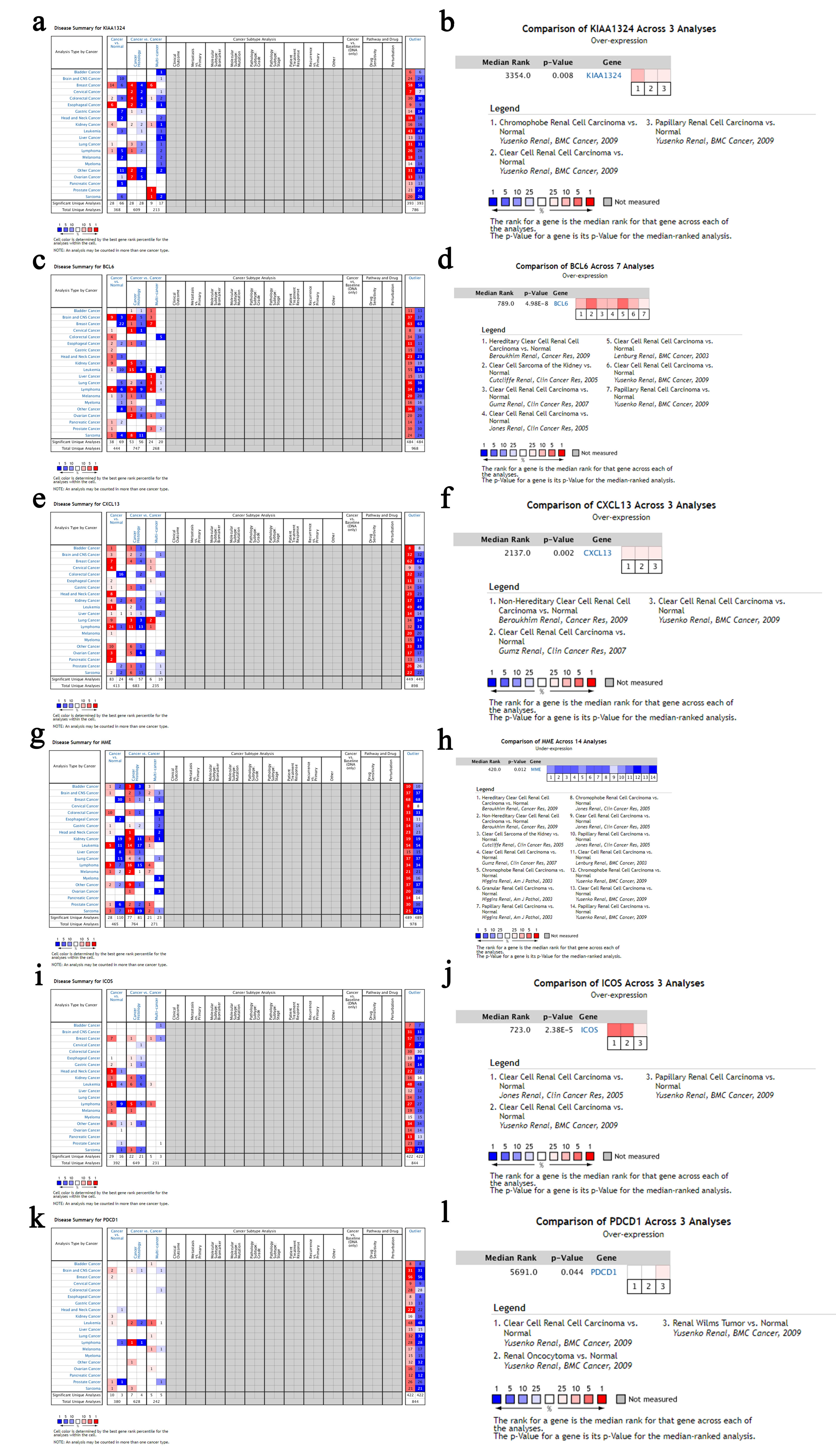

**Figure S6** The expression levels of KIAA1324 and surface markers among different cancer types and multiple studies in the Oncomine database.

(a) and (b): KIAA1324; (c) and (d): BCL6; (e) and (f): CXCL13; (g) and (h): MME; (i) and (j): ICOS; (k) and (l): PDCD1.

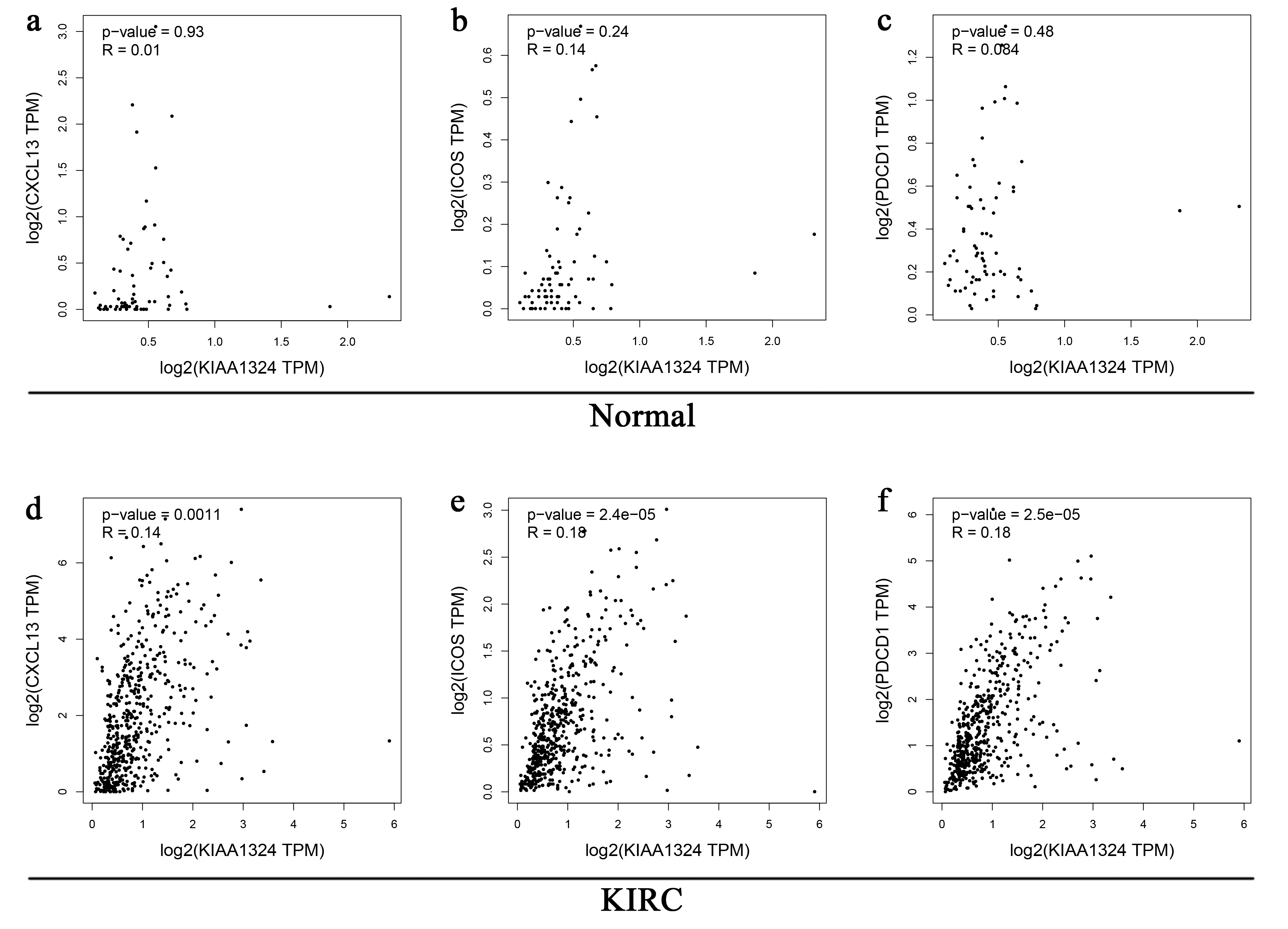

**Figure S7** The correlation of each surface marker and KIAA1324 between normal kidney and KIRC in GEPIA.

(a) and (d): CXCL13; (b) and (e): ICOS; (c) and (f): PDCD1.

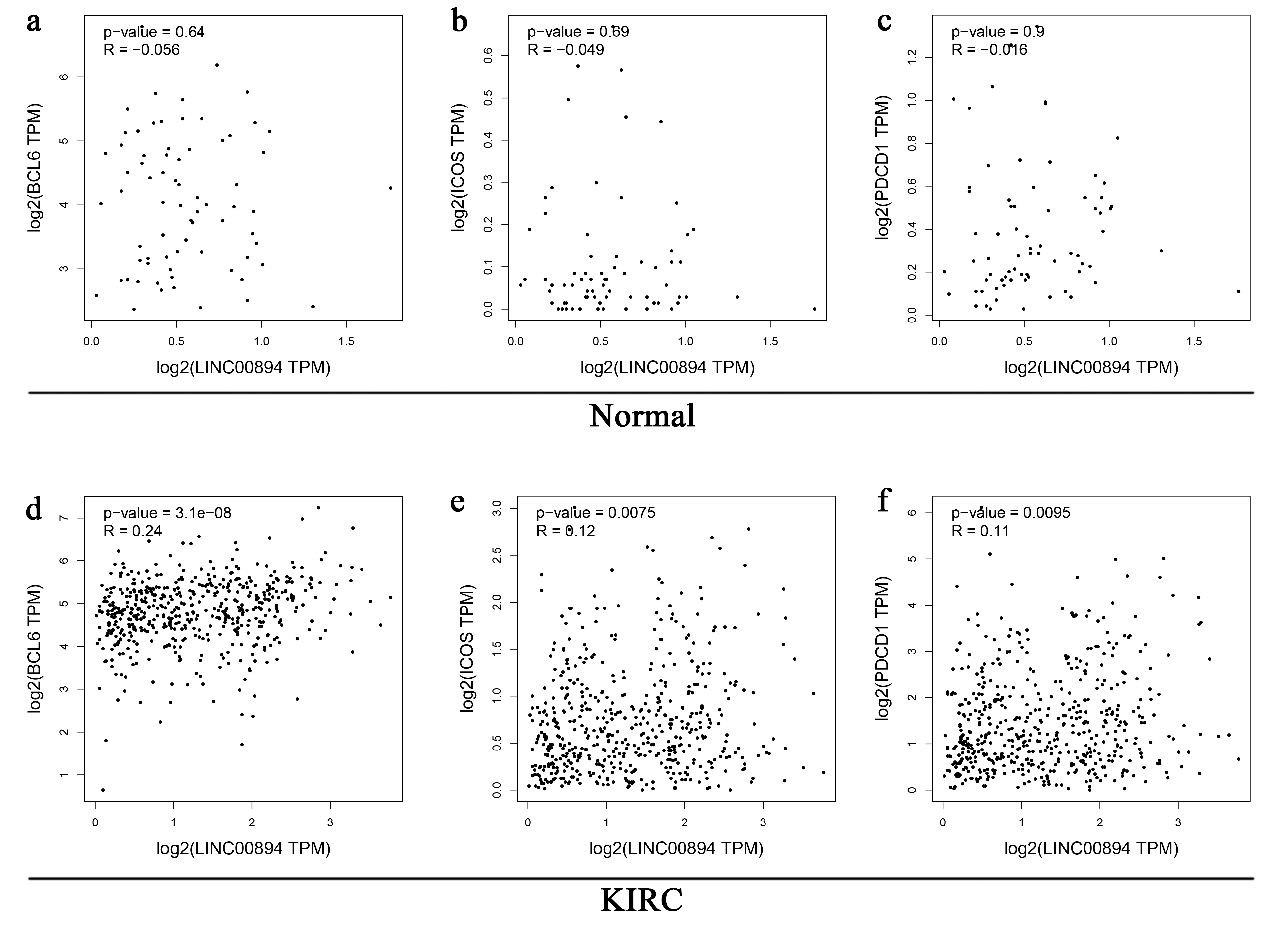

**Figure S8** The correlation of each surface marker and LINC00894 between normal kidney and KIRC in GEPIA.

(a) and (d): BCL6; (b) and (e): ICOS; (c) and (f): PDCD1.

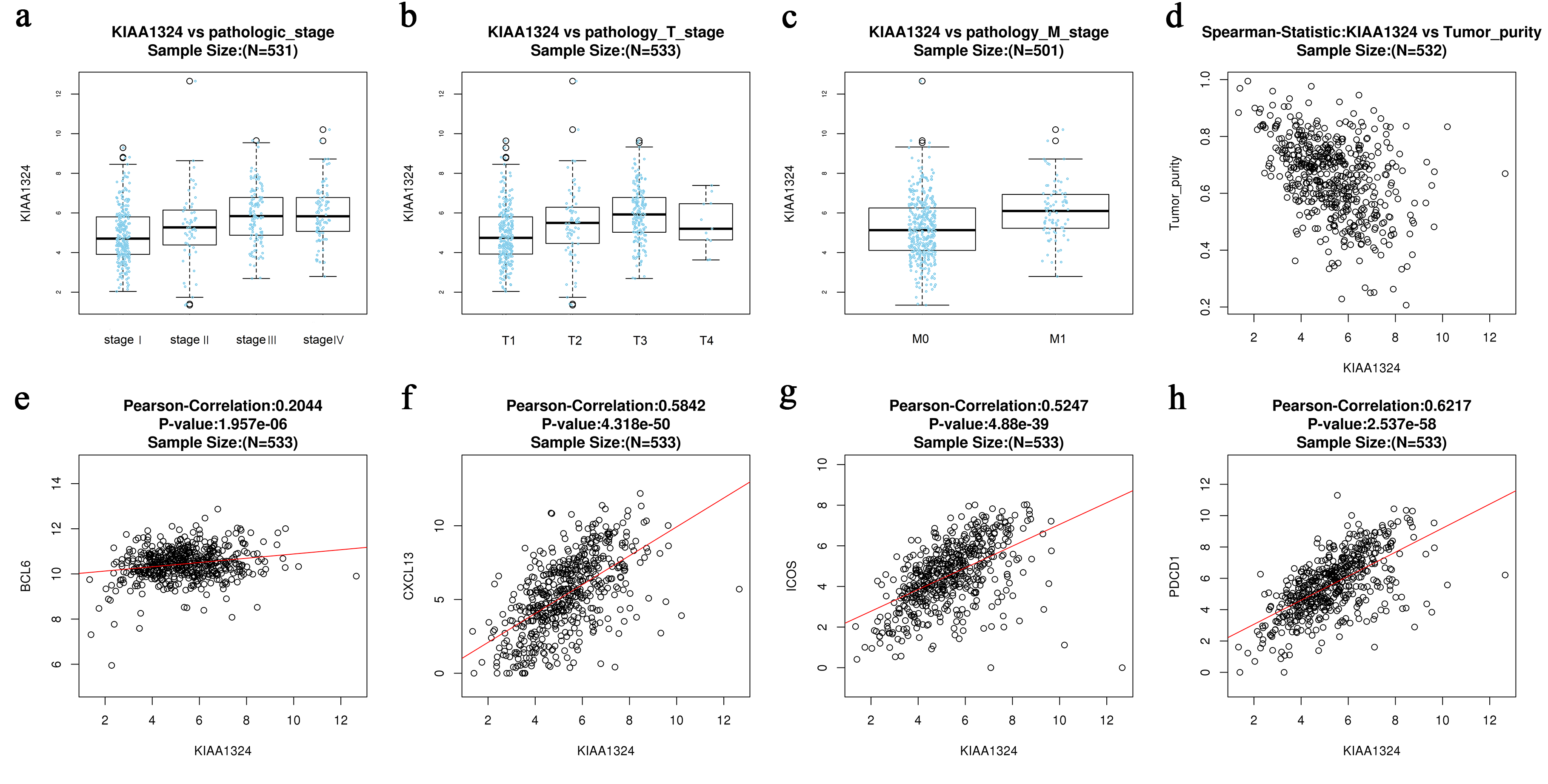

**Figure S9** The validation results of LinkedOmics database.

(a) The expression of KIAA1324 in pathologic stage; (b) The expression of KIAA1324 in T stage; (c) The expression of KIAA1324 in M stage; (d) The correlation of KIAA1324 and tumor purity; (e) The correlation of KIAA1324 and BCL6; (f) The correlation of KIAA1324 and CXCL13; (g) The correlation of KIAA1324 and ICOS; (h) The correlation of KIAA1324 and ICOS.

**Supporting Tables**

**Table S1 Univariate Cox analysis results of ceRNA network**

| **Gene Symbol** | **HR** | **HR.95L** | **HR.95H** | ***P*-value** |
| --- | --- | --- | --- | --- |
| CELSR3 | 1.304119256 | 1.159640751 | 1.466598197 | 9.33E-06 |
| ANLN | 1.329086523 | 1.167770409 | 1.512686888 | 1.64E-05 |
| SH2D2A | 1.244092821 | 1.113326249 | 1.39021868 | 0.000115925 |
| POU2F2 | 1.315843159 | 1.139785949 | 1.519095074 | 0.000180168 |
| TLL1 | 0.799248615 | 0.731422746 | 0.873364073 | 7.32E-07 |
| VPS13D | 0.539004016 | 0.428061528 | 0.678699929 | 1.47E-07 |
| POLQ | 1.359879955 | 1.174278375 | 1.574816952 | 4.03E-05 |
| RELT | 1.928339721 | 1.544934134 | 2.406894896 | 6.40E-09 |
| PRDM1 | 0.784953186 | 0.667454007 | 0.923137023 | 0.003426233 |
| SLC2A3 | 1.159878813 | 1.020924269 | 1.31774599 | 0.022724723 |
| PTHLH | 1.0697084 | 1.011032342 | 1.131789769 | 0.019224168 |
| DTX2 | 1.879154642 | 1.430535346 | 2.468462019 | 5.82E-06 |
| SEL1L3 | 0.868958073 | 0.772108231 | 0.97795633 | 0.019823645 |
| MYO9B | 1.394636541 | 1.02724158 | 1.893431028 | 0.032986137 |
| FAM118A | 1.311823276 | 1.099181287 | 1.565601897 | 0.002629851 |
| CAPN15 | 1.325846449 | 1.021821397 | 1.720328828 | 0.033804034 |
| NDRG1 | 0.765478534 | 0.673119352 | 0.870510384 | 4.62E-05 |
| KCNN4 | 1.5239899 | 1.339718232 | 1.733607233 | 1.48E-10 |
| SNRNP70 | 1.668850655 | 1.344346252 | 2.071685404 | 3.45E-06 |
| GRIN2D | 1.350296307 | 1.20113264 | 1.51798399 | 4.94E-07 |
| GRB10 | 0.58601179 | 0.498295172 | 0.689169467 | 1.05E-10 |
| EZH2 | 1.788604972 | 1.424552204 | 2.245693585 | 5.51E-07 |
| UNC5B | 0.842901012 | 0.73403986 | 0.967906724 | 0.015423084 |
| RASD1 | 0.890005761 | 0.80360735 | 0.985693144 | 0.025316235 |
| KAT2A | 1.857658357 | 1.549550855 | 2.227028922 | 2.18E-11 |
| CNTNAP1 | 1.748133223 | 1.474482526 | 2.072571028 | 1.27E-10 |
| USP46 | 0.717808244 | 0.550075093 | 0.936687885 | 0.01462151 |
| CCND1 | 0.750917487 | 0.663221924 | 0.850208734 | 6.15E-06 |
| APBB3 | 1.4610258 | 1.223890991 | 1.744106628 | 2.72E-05 |
| NPHP3 | 1.803904185 | 1.445077364 | 2.251831209 | 1.85E-07 |
| UXS1 | 0.710448812 | 0.530776287 | 0.950942096 | 0.021555193 |
| KIAA1324 | 1.26323149 | 1.144049202 | 1.394829692 | 3.81E-06 |
| SIPA1L2 | 0.709428169 | 0.59154655 | 0.850800883 | 0.000213329 |
| RSRP1 | 1.298586094 | 1.116387832 | 1.510519728 | 0.000705717 |
| PLAGL1 | 1.17209649 | 1.003030672 | 1.369659194 | 0.045712322 |
| TNFRSF10B | 1.429284407 | 1.095764851 | 1.864317801 | 0.00842564 |
| CDKN2C | 1.285332596 | 1.054808739 | 1.566236438 | 0.012807674 |
| PREX1 | 0.653303464 | 0.548862108 | 0.777618658 | 1.67E-06 |
| RUNX2 | 1.376853251 | 1.171703431 | 1.6179221 | 0.000102372 |
| SIX1 | 1.206735848 | 1.059463576 | 1.374479915 | 0.004657916 |
| EGLN3 | 0.91793829 | 0.842697512 | 0.999896989 | 0.049724673 |
| KMT5C | 1.903875957 | 1.569624641 | 2.309306037 | 6.28E-11 |
| NHSL1 | 0.792056545 | 0.656661854 | 0.95536777 | 0.014795933 |
| FHOD1 | 1.473902669 | 1.118581381 | 1.942093009 | 0.00584822 |
| LIMD2 | 1.35654594 | 1.165030226 | 1.579544331 | 8.60E-05 |
| BRIP1 | 1.220530167 | 1.020330681 | 1.460010873 | 0.029246535 |
| IRF4 | 1.115975959 | 1.009520564 | 1.23365723 | 0.031935997 |
| DNA2 | 1.548403768 | 1.317710493 | 1.819484812 | 1.09E-07 |
| HECW2 | 0.745994157 | 0.671725273 | 0.828474534 | 4.33E-08 |
| MAP3K12 | 1.927372607 | 1.552899823 | 2.392147332 | 2.63E-09 |
| SLC25A37 | 1.794296881 | 1.483598652 | 2.170062161 | 1.68E-09 |
| TRIM36 | 1.326495501 | 1.140993604 | 1.54215616 | 0.000236841 |
| BMP6 | 0.785786328 | 0.701890239 | 0.879710415 | 2.86E-05 |
| SLC25A27 | 1.140838593 | 1.020942294 | 1.274815142 | 0.020028885 |
| TSC22D3 | 0.823397919 | 0.712355113 | 0.951750215 | 0.008562448 |
| FBXO41 | 1.204719692 | 1.083582096 | 1.339399701 | 0.000571963 |
| INTS6L | 1.481780712 | 1.200854393 | 1.828426569 | 0.000245867 |
| OTOGL | 0.776486566 | 0.699341093 | 0.862142084 | 2.15E-06 |
| VKORC1 | 1.395383412 | 1.148888572 | 1.694763891 | 0.000780843 |
| KIFC2 | 1.396967242 | 1.196579395 | 1.630913488 | 2.32E-05 |
| INSR | 0.640239462 | 0.551084345 | 0.743818206 | 5.60E-09 |
| NETO2 | 0.856518012 | 0.755617383 | 0.970892308 | 0.015440398 |
| CLCN5 | 0.663226602 | 0.586143883 | 0.750446329 | 7.31E-11 |
| APLN | 0.859607897 | 0.764030649 | 0.967141485 | 0.01188546 |
| PHLDA3 | 1.194127002 | 1.018652687 | 1.399828731 | 0.028679217 |
| ATAD5 | 1.598154314 | 1.277581971 | 1.99916504 | 4.05E-05 |
| BDNF | 0.902876133 | 0.832353181 | 0.979374298 | 0.013807701 |
| C8orf4 | 0.833035858 | 0.730605185 | 0.949827287 | 0.006354261 |
| AURKB | 1.589967667 | 1.408089863 | 1.795337959 | 7.34E-14 |
| RFLNB | 0.768117096 | 0.662725625 | 0.890268689 | 0.000459027 |
| TSPYL2 | 1.274859286 | 1.072852267 | 1.514902143 | 0.005799864 |
| OLFML2A | 0.832871592 | 0.74529609 | 0.930737593 | 0.001254223 |
| MYBL1 | 1.314258365 | 1.131832096 | 1.526087708 | 0.000338115 |
| ZNF395 | 0.771759774 | 0.671088528 | 0.887532901 | 0.000280155 |
| SPRY4 | 0.781596304 | 0.678237078 | 0.900706851 | 0.000661674 |
| AC005154.1 | 1.47686845 | 1.254655314 | 1.738437956 | 2.78E-06 |
| TCF4 | 0.750433675 | 0.651765314 | 0.864039077 | 6.56E-05 |
| ANKRD13B | 1.461043679 | 1.237996305 | 1.724277062 | 7.26E-06 |
| MXD3 | 1.712793376 | 1.48215516 | 1.979321213 | 3.04E-13 |
| EPB41L4A-AS1 | 0.734679771 | 0.613094237 | 0.88037749 | 0.000837158 |
| LINC00894 | 1.304107159 | 1.169710083 | 1.453946159 | 1.71E-06 |
| LINC00893 | 1.222410577 | 1.097606589 | 1.361405475 | 0.000257255 |
| NEAT1 | 1.167841707 | 1.041181883 | 1.309909704 | 0.008074001 |
| PVT1 | 1.590469853 | 1.309435025 | 1.931821209 | 2.90E-06 |
| MALAT1 | 1.251768161 | 1.108630044 | 1.413387214 | 0.000289582 |
| SNHG1 | 1.434495996 | 1.174170085 | 1.752538911 | 0.000413274 |
| AC015813.1 | 1.351878981 | 1.18609556 | 1.540834347 | 6.28E-06 |
| AC016876.2 | 1.318850091 | 1.089335061 | 1.596722281 | 0.004552003 |
| hsa-let-7e-5p | 0.775536345 | 0.639665508 | 0.940267396 | 0.009689872 |
| hsa-miR-125a-5p | 0.654743452 | 0.526949561 | 0.813529453 | 0.000131969 |
| hsa-miR-130b-3p | 1.821678094 | 1.529463672 | 2.169722066 | 1.78E-11 |
| hsa-miR-181b-5p | 0.811242423 | 0.665356447 | 0.989115342 | 0.038624374 |
| hsa-miR-200b-3p | 0.869726087 | 0.762565678 | 0.991945334 | 0.037478626 |
| hsa-miR-204-5p | 0.895615097 | 0.847500083 | 0.94646174 | 9.12E-05 |
| hsa-miR-21-5p | 1.716063619 | 1.403080292 | 2.098863738 | 1.47E-07 |
| hsa-miR-342-3p | 1.376362734 | 1.113312246 | 1.701566099 | 0.003158861 |
| hsa-miR-590-3p | 1.449787696 | 1.161439627 | 1.809723309 | 0.00102813 |

**Table S2 Univariate Cox analysis results of immune cells**

| **Cell type** | **HR** | **HR.95L** | **HR.95H** | ***P*-value** |
| --- | --- | --- | --- | --- |
| T cells follicular helper | 505230.5082 | 11.79309815 | 21644682613 | 0.015803535 |
| Mast cells resting | 3.45E-06 | 1.48E-10 | 0.080730352 | 0.014273779 |

**Table S3 The comparison results in the Oncomine database**

| **Gene** | **Median rank** | ***P*-value** |
| --- | --- | --- |
| KIAA1324 | 3354.0 | 0.008 |
| BCL6 | 789.0 | 4.98E-8 |
| CXCL13 | 2137.0 | 0.002 |
| MME | 420.0 | 0.012 |
| ICOS | 723.0 | 2.38E-5 |
| PDCD1 | 5691.0 | 0.044 |
